# Examining the status of improved air quality due to COVID-19 lockdown and an associated reduction in anthropogenic emissions

**DOI:** 10.1101/2020.08.20.20177949

**Authors:** Srikanta Sannigrahi, Anna Molter, Prashant Kumar, Qi Zhang, Bidroha Basu, Arunima Sarkar Basu, Francesco Pilla

## Abstract

Clean air is a fundamental necessity for human health and well-being. The COVID-19 lockdown worldwide resulted in controls on anthropogenic emission that have a significant synergistic effect on air quality ecosystem services (ESs). This study utilised both satellite and surface monitored measurements to estimate air pollution for 20 cities across the world. Sentinel-5 Precursor TROPOspheric Monitoring Instrument (TROPOMI) data were used for evaluating tropospheric air quality status during the lockdown period. Surface measurement data were retrieved from the Environmental Protection Agency (EPA, USA) for a more explicit assessment of air quality ESs. Google Earth Engine TROPOMI application was utilised for a time series assessment of air pollution during the lockdown (1 Feb to 11 May 2020) compared with the lockdown equivalent periods (1 Feb to 11 May 2019). The economic valuation for air pollution reduction services was measured using two approaches: (1) median externality value coefficient approach; and (2) public health burden approach. Human mobility data from Apple (for city-scale) and Google (for country scale) was used for examining the connection between human interferences on air quality ESs. Using satellite data, the spatial and temporal concentration of four major pollutants such as nitrogen dioxide (NO_2_), sulfur dioxide (SO_2_), carbon monoxide (CO) and the aerosol index (AI) were measured. For NO_2_, the highest reduction was found in Paris (46%), followed by Detroit (40%), Milan (37%), Turin (37%), Frankfurt (36%), Philadelphia (34%), London (34%), and Madrid (34%), respectively. At the same time, a comparably lower reduction of NO_2_ is observed in Los Angeles (11%), Sao Paulo (17%), Antwerp (24%), Tehran (25%), and Rotterdam (27%), during the lockdown period. Using the adjusted value coefficients, the economic value of the air quality ESs was calculated for different pollutants. Using the public health burden valuation method, the highest economic benefits due to the reduced anthropogenic emission (for NO2) was estimated in US$ for New York (501M $), followed by London (375M $), Chicago (137M $), Paris (124M $), Madrid (90M $), Philadelphia (89M $), Milan (78M $), Cologne (67M $), Los Angeles (67M $), Frankfurt (52M $), Turin (45M $), Detroit (43M $), Barcelona (41M $), Sao Paulo (40M $), Tehran (37M $), Denver (30M $), Antwerp (16M $), Utrecht (14 million $), Brussels (9 million $), Rotterdam (9 million $), respectively. In this study, the public health burden and median externality valuation approaches were adopted for the economic valuation and subsequent interpretation. This one dimension and linear valuation may not be able to track the overall economic impact of air pollution on human welfare. Therefore, research that broadens the scope of valuation in environmental capitals needs to be initiated for exploring the importance of proper monetary valuation in natural capital accounting.

## 1. Introduction

As per the Ecosystem Services (ESs) definition of Millennium Ecosystem Assessment (MA, 2005), provision of clean air is one of the fundamental needs of human lives, which mainly comes from natural vegetation and appropriates by human interferences (Schirpke et al., 2014; Ash et al., 2010; Charles et al., 2020; Baró et al., 2014). The accelerated increases of air pollution across the world that mainly comes from transport emissions, industrial emission, domestic emission, and waste incineration is the primary reason for the degrading status of air quality ecosystem services. The high concentration of air pollutants, including nitrogen dioxide (NO_2_), carbon monoxide (CO), particulate matter (PM_2.5_ and PM_10_), sulfur dioxide (SO_2_), which goes beyond the normal absorption capacity by the green canopy, leading to a paramount impact on the quality of human life (Nowak, 1994; Escobedo et al., 2008; De Carvalho and Szlafsztein, 2019; Gómez-Baggethun and Barton, 2013). The COVID-19 pandemic and its associated restriction on human activities cut down the pollution level drastically across the scale (Kumar et al., 2020a,b; Mahato et al., 2020). Many scholarly works appear on time to discuss the positive effect of COVID-19 lockdown on air quality (Venter et al., 2020, Kumar et al., 2020a; Ogen, 2020; Sasidharan et al., 2020; Sharma et al., 2020). However, a thorough evaluation is needed to measure the synergistic effects of these interventions on air quality ecosystem services.

Air pollution has been reduced drastically due to COVID-19 led lockdown and its resultant restrictions on human activities. Veneter et al. (2020) had examined both tropospheric and ground air pollution levels using satellite data and a network of > 10,000 air quality stations across the world and found that 29% reduction of NO_2_ (with 95% confidence interval –44% to –13%), 11% reduction of Ozone (O3), and 9% reduction of PM_2.5_ during the first two weeks of lockdown (Venter et al., 2020). Kerimray et al. (2020) study at Almaty, Kazakhstan, found that the effect of city-scale lockdown, which was effective on March 19, 2020, has resulted in 21% reduction of PM_2.5_ with spatial variation of 6 – 34%. The CO (49% reduction) and NO_2_ (35% reduction) concentration has also been reduced substantially. In the same period, an increase (15%) in O3 levels is also observed in Almaty, Kazakhstan (Kerimray et al., 2020). Mahato et al. (2020) had reported a sharp reduction in air pollution in Delhi, one of the most polluted cities in the world. The author found that the concentration of PM_10_ and PM_2.5_ in Delhi was reduced to 60% and 39%, compared to the air pollution levels in 2019 (considered the lockdown period only). The concentration of other pollutants, such as NO_2_ (−52.68%) and CO (−30.35%), have also been reduced substantially during the lockdown period. In addition to this, Mahato et al. study has observed a 40% to 50% improvement in air quality in Delhi within the first week of lockdown. Bao and Zhang, (2020) study combined air pollution and Intracity Migration Index (IMI) data for 44 cities in northern China and found that restriction on human mobility is strongly associated with the reduction of air pollution in these cities. The author found that the air quality index (AQI) in these cities is decreased by 7.80%, as the concentration of five key air pollutants, i.e., SO_2_, PM_2.5_, PM_10_, NO_2_, and CO have decreased by 6.76%, 5.93%, 13.66%, 24.67%, and 4.58%, respectively. Sicard et al., (2020) had observed that due to lockdown and resulted in the restriction on human activities, NO_2_ mean concentrations were reduced substantially in all European cities, which was ∼53% at urban stations. During the same period, the mean concentrations of O3was reported to be increased at the urban stations in Europe, i.e., 24% increases in Nice, 14% increases in Rome, 27% increases in Turin, 2.4% increases in Valencia and 36% in increases in Wuhan (China). Otmani et al., (2020) study at Morocco using three-dimensional air mass backward trajectories and HYSPLIT model found that PM_10_, SO_2_, and NO_2_ are reduced up to 75%, 49%, and 96% during the lockdown period. In the southeast Asian (SEA) countries, (Kanniah et al., 2020) study found that PM_10_, PM_2.5_, NO_2_, SO_2_, and CO concentrations have been decreased by 26–31%, 23–32%, 63–64%, 9–20%, and 25–31% during the lockdown period in Malaysia. Kumar et al., (2020a) examined the impacts of COVID-19 mitigation measures on the reduction of PM_2.5_ in five Indian cities (Chennai, Delhi, Hyderabad, Kolkata, and Mumbai), using in-situ measurements from 2015 to 2020. Kumar et al. study found that during the lockdown period (25 March to 11 May), the PM_2.5_ concentration in the selected cities has been reduced by 19 to 43% (Chennai), 41–53% (Delhi), 26–54% (Hyderabad), 24–36% (Kolkata), and 10–39% (Mumbai), respectively. This study also found that cities with higher traffic volume exhibited a greater reduction of PM_2.5_.

The level of air pollution has a severe impact on human health and overall well-being. Air pollution is responsible for nearly 5 million deaths each year globally (IHME, 2020). In 2017, air pollution had contributed to 9% of deaths, ranges from 2% in the high developed country to a maximum 15% in low-developed countries, especially in South and East Asia (IHME, 2020). Based on Disability-Adjusted Life Years (DALYs) statistics, which demonstrate of losing one year of good health due to either premature mortality or disability caused by any factors, it has been estimated that air pollution is the 5^th^ largest contributor to overall disease burden, only after high blood pressure, smoking, high blood sugar, and obesity, respectively. The adverse impact of air pollution on human health is not only limited to (low)developing countries. In the European regions, nearly 193,000 deaths in 2012 were attributed to airborne particulate matter (Ortiz et al., 2017). In addition, it has been found that air pollution in China is accountable for 4000 deaths each day, i.e., 1.6 million casualties in 2016 (Rohde and Muller, 2015; Wang and Hao, 2012). By looking at the adverse effects of air pollution on COVID-19 counts, Chen et al., (2020) found that reduction in PM_2_·5 during the lockdown period helped to avoid a total of 3214 PM_2_·5 related deaths (95% CI 2340–4087). Chen et al., (2020) also estimated that COVID-19 lockdown and resulted cut down of air pollution brought multi-faceted health benefits to non-COVID mortalities. Several research studies (He et al., 2020; L. et al., 2015; Dutheil et al., 2020a) have echoed the surmountable effects of air pollutants on human lives and found that an increase in 10μg m^−3^ of NO_2_ per day will be responsible for a 0.13% increases of all-cause mortality (He et al., 2020). The mortality rate would be around 2% when the 5-day NO_2_ level would reach 10μg m^−3^(Monica et al., 2011). In addition to this, L. et al. (2015) estimated that the increase in 8.1 ppb in NO_2_ is attributed to 1.052 increases in global hazard ratio related to air pollution.

Ecosystem Services (ESs) are the supports and benefits (*provisioning*, such as food and water; *regulating* such as management of floods, drought, land degradation, and disease; supporting such as soil formation and nutrient cycling; and *cultural* such as recreational, spiritual, religious and other non-material) that humans have free access from natural environment and ecosystems, which adds to human well-being (Fisher et al., 2009; Costanza et al., 1997; Braat and de Groot, 2012; Sannigrahi et al., 2018; Sannigrahi et al., 2019). The ecosystem service value (ESV) is a comprehensive assessment and has proven to be an alternative appraisal between environment and human development for sustainable natural resource management (Braat and de Groot, 2012; Potschin and Haines-Young, 2013; Pandeya et al., 2016; Sannigrahi et al., 2020c; Sannigrahi et al., 2020b; Adekola et al., 2015). The growing importance of ESs helps in adjusting the cost-benefit analysis by evaluating both the negative and positive effects of human interferences on the natural environment and ecosystems. Considering the plausible application of ecosystem service valuation in different strata of planning, priorities should be given to developing a suitable valuation framework for estimating the biophysical and economic values of the key ESs (Bastian et al., 2013; Burkhard et al., 2014; Spangenberg et al., 2014; Affek and Kowalska, 2017; Sannigrahi et al., 2019). Due to unawareness about the importance of ESs on natural capital formation and human well-being, the ecosystem service valuation research was neglected for an extended period (Jack et al., 2008). To overcome this, several national and international valuation framework were formed, including The Economics of Ecosystems and Biodiversity (TEEB), The Inter-governmental Science-Policy Platform on Biodiversity and Ecosystem Services (IPBES), Millennium Ecosystem Assessment (MA 2005), Ecosystem Service Partnership (ESP) to name a few (Burkhard et al., 2009; Costanza et al., 2014; Comberti et al., 2015).

It is now well-established by many data-driven experiments that the accelerated rate of air pollution can have a substantial impact on overall human well-being. Due to this pandemic, the world witnessed an extraordinary transformation in all strata of lives, such as adopting digital alternatives to carry out the routine life and imposing national scale lockdown to restrict human mobility and social activity, to prevent the spread of infection. Additionally, as it is observed by many studies across the scale, the long term restriction on human mobility resulted in the reduction of road traffic, which improved the air quality status of a region. The importance of this human-induced reduction of air pollution needs to be evaluated in a way so that the same could be used as a reference for future decision making and policy formation. The present research thus made an effort to investigate the human impact on the natural environment by taking COVID-19 lockdown and its resultant effects of air pollution as a case for the experiment. The economic valuation was carried out to assess the synergistic effect of this pandemic on air pollutions at 20 cities across the world. The main objectives of this study are: (1) to estimate the spatiotemporal changes in air pollution during 1 February to 11 May in 2019 and 2020 using both satellite and ground monitoring data; (2) to estimate the air quality ecosystem service using multiple economic valuation approaches; (3) to evaluate the association between human mobility and reduction of air pollution.

## 2. Materials and methods

### 2.1 Data source and data preparation

A total of 20 cities have been selected for evaluating the effect of lockdown on air quality ESs. These cities are Antwerp, Barcelona, Brussels, Chicago, Cologne, Denver, Frankfurt, London, Los Angeles, Madrid, Milan, New York, Paris, Philadelphia, Rotterdam, Sao Paulo, Tehran, Turin, and Utrecht. These cities have been considered based on two criteria: high air pollution and high COVID-19 casualties. Most of the cities listed here are from European and American countries. These countries reported more COVID-19 casualties compared with the Asian and Latin American countries (as of 11 May 2020) (WHO, 2020; Sannigrahi et al., 2020a). Sentinel 5P time series pollution data were also used to identify the most polluted cities. Both satellite and ground air pollution data were utilised for evaluating the positive effects of lockdown on the air quality index of these cities. For comparison, the satellite-based air pollution was measured from 01 February to 11 May for both 2019 (lockdown equivalent period) and 2020 (lockdown period). The concentration of four key air pollutants, nitrogen dioxide (NO_2_), sulfur dioxide (SO_2_), carbon monoxide (CO), and aerosol index (AI) concentration, was computed for both 2019 and 2020 using Sentinel 5P data. For six cities, i.e., Chicago, Denver, Detroit, Los Angeles, New York, and Philadelphia, the ground monitored air pollution data was collected for a more explicit assessment of air quality ESs. However, the ground monitored data was not adequate for the spatial evaluation for most of the cities considered in this study. Therefore, the in-situ data was only used for time series assessment of air pollutions, and the satellite measured pollution estimates were utilised for the spatially explicit appraisal and economic valuation. Human mobility data, including driving and transit for the selected cities, were collected from Apple (for city-scale) and Google (for country scale) mobility reports. In addition to this, the gridded human settlement data and population density data (pixel format) were collected from the Socio-Economic Data Application Center, National Aeronautics and Space Application data center (SEDAC, NASA). For evaluating the total air pollution reduction of these 20 cities in a more accurate way, the Geographical Information System (GIS) enabled city boundary (shapefile format) was extracted from the OpenStreetMap (OSM) application. Two consecutive steps were followed to get the boundary of these cities. First, the OSM relation identifier number (OSM id) was generated for all the 20 cities using Nominatim, a search engine for OpenStreetMap data. Then, the OSM relation id of each city was ingested in the OSM polygon creation application interface, which generates the geometry (both actual and simplified) of the relation id in poly, GeoJSON, WKT or image formats. The formatted image geometry of the cities was then imported in ArcGIS Pro software, and the city boundary was extracted using an automatic digitisation function.

### 2.3 Estimation of air pollution

#### 2.3.1 Sentinel 5P TROPOMI data and TROPOMI Explorer Application

The ESA (European Space Agency) Sentinel-5 Precursor (S 5P) is an example of low earth Sun-synchronous Orbit (SSO) polar satellite that provides information of tropospheric air quality, climate dynamics and ozone layer concentration for the time period 2015–2022 (Veefkind et al., 2012). The ESA led S 5P mission is one of the few missions that is intended to measure air and climatic variability from the space-borne application. The S 5P mission is associated with the Global Monitoring of the Environment and Security (GMES) space programme. The TROPOspheric Monitoring Instrument (TROPOMI) payload of S 5P mission was designed to measure the tropospheric concentration of few key air pollutants, i.e., ozone (O3), NO_2_, SO_2_, CO, CH_4_, CH_2_O and aerosol properties in line with Ozone Monitoring Instrument (OMI) and SCanning Imaging Absorption spectroMeter for Atmospheric CartograpHY (SCIAMACHY) programme (Veefkind et al., 2012). TROPOMI measures the concentration of key tropospheric constituents at 7 × 7 km^2^ spatial unit. This default spatial scale was downscaled into 1km × 1km scale for city-scale analysis and subsequent interpretation. In this study, the spatial and temporal variability of four key air pollutants was extracted and mapped from the TROPOMI measurements using the Google Earth Engine cloud platform. For this purpose, an interactive application called TROPOMI Explorer App, developed by Google developers teams (Google, 2020; Braaten, 2020), was utilised to facilitate quick and easy S5P data exploration and to examine the changes in air pollution in both cross-sectional and longitudinal way. Spatial visualisation and time series charts for the selected air pollutants were also prepared with the help of this TROPOMI Explorer application. The other accessories of this application, such as NO_2_ time series inspector, NO_2_ temporal comparison, NO_2_ time-series animation, were also utilised for different computational purposes.

#### 2.3.2 Ground pollution data

Ground monitored air quality data was available only for a few cities considered in the study, including Chicago, Denver, Detroit, Los Angeles, New York, and Philadelphia. Thus, these cities were selected for the ground data-driven analysis. Ground monitored data for these cities were collected from the U.S. Environmental Protection Agency (US EPA). This data is available for a daily scale and for six key pollutants, such as CO, NO_2_, O_3_, PM_2.5_, PM_10_, and SO_2_, respectively. The in-situ air pollution concentration at a daily scale was considered only for the time series assessment of pollution concentration. Additionally, the said in-situ data had not been used for any validation and calibration of satellite pollution estimates. The time series (2000–2020) air quality index (AQI) of these selected cities were also generated using the multilayer time plot function. The overall AQI values were sub-divided into six groups, i.e., good, moderate, unhealthy for sensitive population groups, unhealthy, very unhealthy, and hazardous, respectively. In addition to this, the single year AQI data was also extracted for the selected cities from the EPA. The number of unhealthy days for each pollutant was measured using the EPA AQI plot function. The combination of two different pollutants, such as CO and NO_2_, PM_10_ and PM_2.5_, was permuted to assess the yearly AQI status of the cities. As several studies reported the increment of O3 due to the reduction of GHG emissions, this study also evaluated the O3 exceedances for the current year compared to the average O3 concentration of the last 5 and 20 years. This particular task was implemented using the EPA Ozone exceedances plot function (EPA, 2020). **Table. S1** provides the criteria of categorisation for each index.

### 2.4 Environmental significance of improving air quality status

The accelerating increases of air pollution in cities is a major concern across the world (Chan and Yao, 2008; Kim Oanh et al., 2006; Mayer, 1999; Guttikunda et al., 2014; Abhijith et al., 2017; Rai et al., 2017; Pilla and Broderick, 2015). Various policies have been implemented for managing the city-based air pollution that mainly originated from anthropogenic activities from specific sources and sectors (Kumar et al., 2015; Kumar et al., 2016; Baró et al., 2014; Feng and Liao, 2016; Zhang et al., 2016). These include the Directive 2010/75/EU on industrial emissions, initiated by European Commission to define ‘‘Euro standards’’ for measuring the road vehicle emissions and the Directive 94/63/EC for calculating volatile organic compounds emissions from petrol storage (Baro et al., 2014). The reduction of these gaseous pollutants by green canopy has significant economic importance (Kumar et al., 2019). Two main ecosystem services, such as air quality regulation and climate/gas regulation, are mainly associated with air quality ecosystem services. Several studies have calculated the economic values of NO_2_, SO_2_, CO reductions using various valuation approaches such as carbon tax, the social cost of carbon, shadow price method, marginal cost method, etc. (Guerriero et al., 2016; Castro et al., 2017; Jeanjean et al., 2017; Bherwani et al., 2020). In this study, multiple relevant approaches were adopted for calculating the economic values of the NO_2_, SO_2_, CO, aerosol reduction to gauge the economic benefits of these functions. Since this study has considered the air pollution reduction at the city scale, the public health burden and mean externality valuation approaches were utilised for estimating economic damage due to air pollution and to calculate the economic values of air quality services (Baro et al., 2014; Matthews and Lave, 2000). Unit social damage price due to air pollution was estimated for 2020 using the U.S consumer price index (CPI) inflation calculator (U.S Bureau of Labor Statistics, 2020). Additionally, using the most updated price conversion factors, the mean externality values for the key pollutants was estimated as: CO = 956 $ t^−1^, NOx = 5149 $ t^−1^, SO_2_ = 3678 $ t^−1^, PM_10_ = 7907 $ t^−1^.

The public health burden valuation approach has also been utilised for economic valuation of air quality ESs (Kumar et al., 2020a, Etchie et al. 2018; Hu et al., 2015; Sharma et al., 2020; Sahu and Kota, 2017; COMEAP, 2009). The calculation of public health burden and the associated economic burden was conducted by three subsequent steps: first, estimation of population-weighted average concentration; second, estimation of health burden or a number of premature mortality attributable to air pollution; and third, the economic burden due to excess air pollution and economic benefits subject to the reduction of air pollution levels during the lockdown period. The population-weighted average concentration (PWAC) was measured as follows:

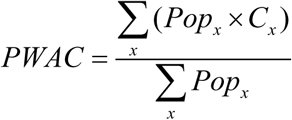

Where *Pop*_*x*_ is the population count of a pixel, *C*_*x*_ is the average pollution concentration (1 Feb to 11 May 2020), 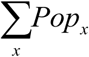 is the total population count of the city, *PWAC* is the population-weighted average concentration. The PWAC was estimated using ArcPy Python module. Gridded population data from SEDAC, NASA, was utilised for this task. Pollution and gridded population data for the same time period were used for estimations of PWAC.

Following, the health burden (HB), which refers premature deaths attributable to short-term exposure to air pollutants was estimated for the study period (1 February to 11 May 2020). The reduction in health burden (ΔHB) was also measured by calculating the difference between the previous and later HB estimates.

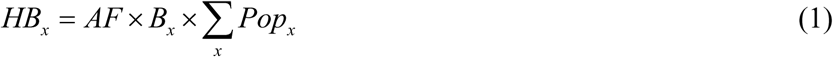

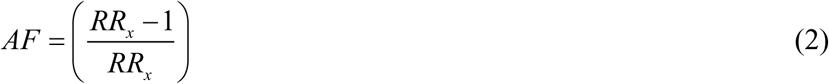

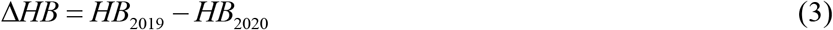

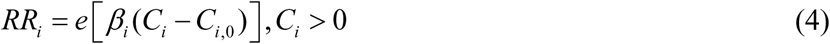

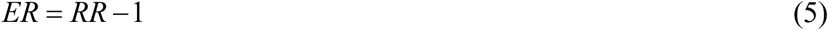

Where *HB*_*x*_ is the health burden of city x, *AF* is the attributable fraction associated with the relative risk of each pollutant, *RR*_*i*_ is the relative risk of pollutant i, *B*_*x*_ is baseline cause-specific mortality rate per 100,000 population. For calculating *B*_*x*_, country-wise cardiovascular and chronic respiratory baseline mortality rate was collected from Global Burden of Disease study of 2017 (IHME, 2020). *Pop*_*x*_ is the population of city x derived from the SEDAC, NASA gridded population count data. Δ*HB* is the difference in health burden (or avoidance of premature death due to the reduction of air pollution) from 1^st^ February to 11^th^ May 2020 compared to the same period in 2019. *HB*_2019_ and *HB*_2020_ is the health burden estimates in 2019 and 2020 (estimated for 1 February to 11 May time period). *β*_*i*_ is the exposure-response relationship coefficient, indicates the excess risk of health burden (such as mortality) per unit increase of pollutants. *β* is calculated 0.038%, 0.032%, 0.081%, 0.13%, and 0.048% per 1 *μg / m*^3^ increases of PM_2.5_, PM_10_, SO_2_, NO_2_, and O_3_, respectively (Hu et al., 2015; Sharma et al., 2020, Kumar et al., 2020a; Chen et al., 2020). *β* is calculated 3.7% per 1 mg/m^3^ increases of CO. *C*_*i*_ is the concentration of pollutant i, *C*_*i*,0_ is the threshold concentration, below which the pollutant exhibits no obvious adverse health effects (i.e., RR = 1).

The economic burden (EB) and economic benefits of the reduced air pollution concentration were estimated using the value of statistical life (VSL) approach (Etchie et al. 2018; Hu et al., 2015). The VSL represents an individual’s willingness to pay for a marginal reduction in risk of dying. The VSL method has been utilised as a standard approach for ecosystem service valuation of non-marketable commodities and is often used for cost-benefit analysis (OECD, 2014; WHO, 2015), ecosystem service studies (Zhang et al., 2018, 2020). The economic benefits due to avoided premature mortality were estimated as follows:

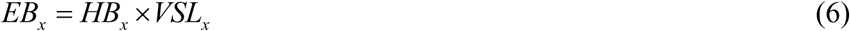

Where *EB*_*x*_ is the economic benefit attributed to the reduction of air pollution and resulted in estimates of avoidable mortality *HB*_*x*_, health burden estimates of city x, *VSL*_*x*_ is the value of statistical life of the country x that corresponds to the city. Using the value transfer method, OECD (2016a) estimated the VSL for the entire world, after incorporating income elasticity beta of 1. Since this study considers cities that cover many diversified economic setup and development background, a uniform income elastic global VSL estimates measured by Viscusi et al., (2017) was considered for the economic valuation and subsequent analysis. As city-specific VSL data is not available for many cities, the VSL estimates for the corresponding countries were taken for the analysis. The 2017 VSL values were converted to 2020 unit price for adjusting price fluctuation. The income adjusted VSL was estimated as Belgium (8 $ millions, used this value for Antwerp and Brussels city), Spain (5 $ millions, this value was used for Barcelona, Madrid), USA (10 $ millions, this value was used for Chicago, Denver, Detroit, Los Angeles, New York, and Philadelphia), Germany (8 $ millions, this was used value for Cologne, Frankfurt), UK (8 $ millions, this value was used for London), Italy (6 $ millions, this value was used for Milan and Turin), France (7 $ millions, this value was used for Paris), Netherlands (9 $ millions, this value was used for Rotterdam and Utrecht), Brazil (2 $ millions, this value was used for Sao Paulo), and Iran (1 $ millions, this value was used for Tehran), respectively (Viscusi et al., 2017) (**Table S5**).

### 2.5 Examining human mobility and its connections with air pollution status

Due to the emergence of COVID-19 pandemic, countries across the world imposed mandatory lockdowns to restrict human-mobility. This reduced motorised traffic, which is one of the key sources of urban air pollution (Chinazzi et al., 2020; De Brouwer et al., 2020). Human mobility could accelerate the transmission of contagious diseases, especially when a larger section of daily commuter uses public transport to maintain their essential daily journey (Sasidharan et al., 2020). Joy et al. and Lara et al. research highlighted a statistically significant association between human mobility that is mainly attributed to public transport and transmissions of acute respiratory infections (ARI) (Troko et al., 2011; Goscé and Johansson, 2018). Joy et al. (2011) also found that the use of public transport during a pandemic outbreak in the UK has increased the risk of ARI infection by six-times. To evaluate the effects of reduced human mobility on air pollution, this study utilised the human mobility data provided by Apple and Google. Apple mobility data includes three mobility components, i.e., driving, walking, and transit (public transport), respectively. The reduction of human mobility during the lockdown period was calculated from the baseline (13 January). Both positive and negative changes in human mobility were recorded in percentage form to eliminate calculation bias and easy comparability across the cities/countries in the world. Among the three mobility components, driving and transit was considered for the evaluation, and walking was discarded from the analysis. Google mobility data was also used in this study which has six components (retail and recreation, grocery and pharmacy, parks, transits, workplace, and residential). This data is available from 15 February 2020 to recent date. Since Google mobility data is not available for city scale, the smallest scale (county/state) was taken for the analysis for which the mobility counts are available. This data is also prepared in percentage format to handle the calculation bias and better understanding of the data.

## 3. Results

### 3.1 Spatial changes in air pollution in different cities due to lockdown

Spatial distribution of four key air pollutants, i.e., NO_2_ (Fig. 1) SO_2_ (Fig. S1), CO (Fig. S2), and aerosol concentration (Fig. S3) is analysed for 20 cities across the world. The spatial distribution of these pollutants was measured from 1 February to May 11 in 2019 and 2020. A sharp reduction in NO_2_ and CO emission is observed for all the cities. This could be due to the lockdown and resultant reduction of transportation and industrial emission. Among the 20 cities, the maximum decrease of NO_2_ concentration is recorded for the European cities, such as Paris, Milan, Madrid, Turin, London, Frankfurt, Cologne, and American cities, such as New York, Philadelphia, etc. (**Fig. 1**). Moreover, among the 20 cities, the highest NO_2_ reduction is recorded in Tehran, and the lowest reduction is found in Los Angeles and Sao Paulo (based on 1^st^ Feb to 11^th^ May pollution data). The SO_2_ emission is evaluated and presented in **Fig. S1**. An incremental trend of SO_2_ emission is observed during the study period. For most cities, SO_2_ concentration was increased during the study period. However, for exceptions, a slight decrease in SO_2_ emission is observed in Rotterdam, Frankfurt, London, and Detroit cities (**Fig. S1**). The spatial distribution of CO is also evaluated using GEE cloud application and Sentinel 5P data and presented in **Fig. S2**. The CO emission is reduced significantly in all the 20 cities. The highest reduction is recorded in Detroit, followed by Barcelona, London, Los Angeles, New York, Philadelphia, Milan, Madrid, etc. (**Fig. 2**). At the same time, CO emission was increased in Cologne, Denver (**Fig. S2**). The spatial distribution of aerosol concentration is also calculated and presented in **Fig. S3**. Aerosol concentration is also found to be decreased during the COVID lockdown with restricted human activities.

**Fig. 1.**
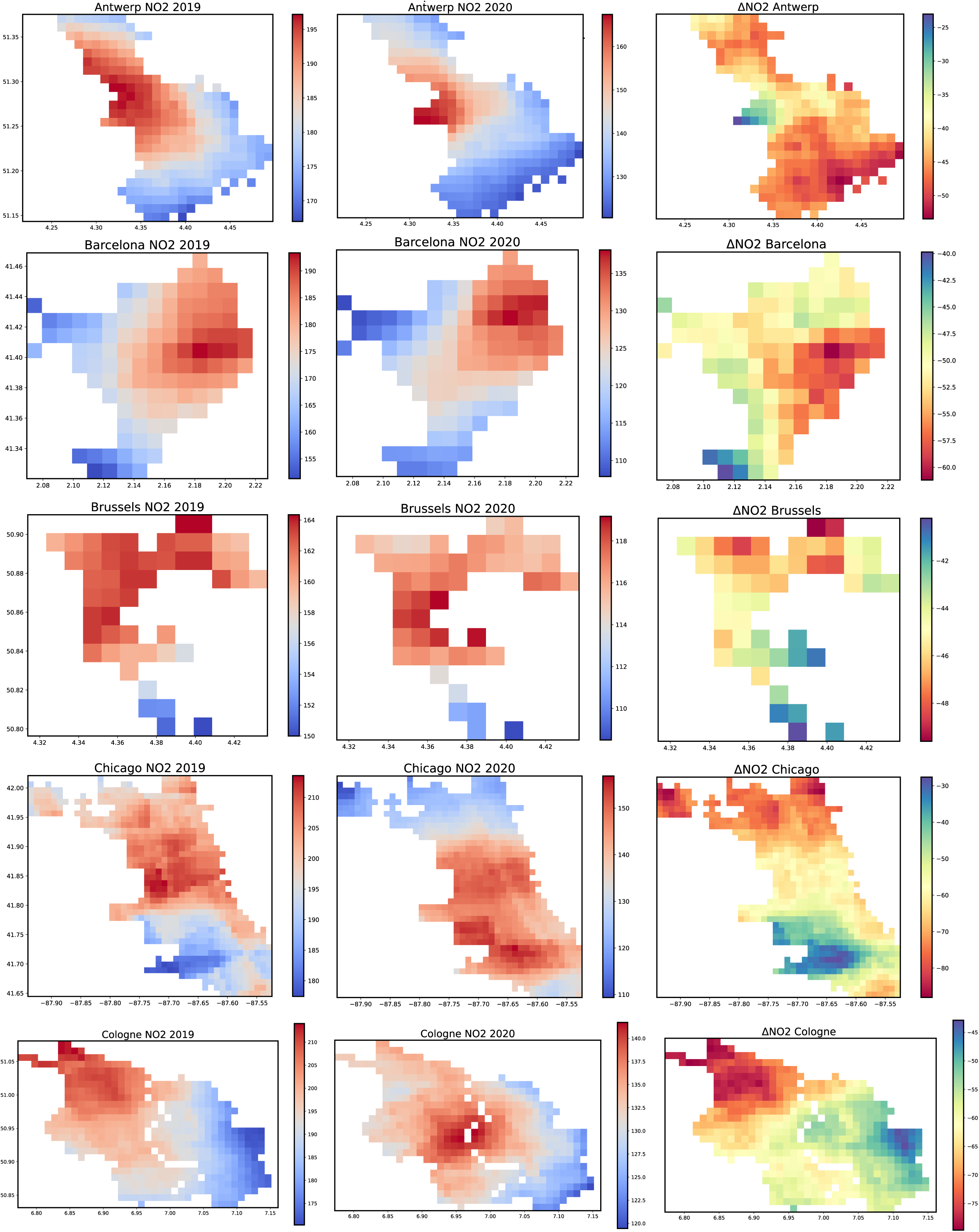
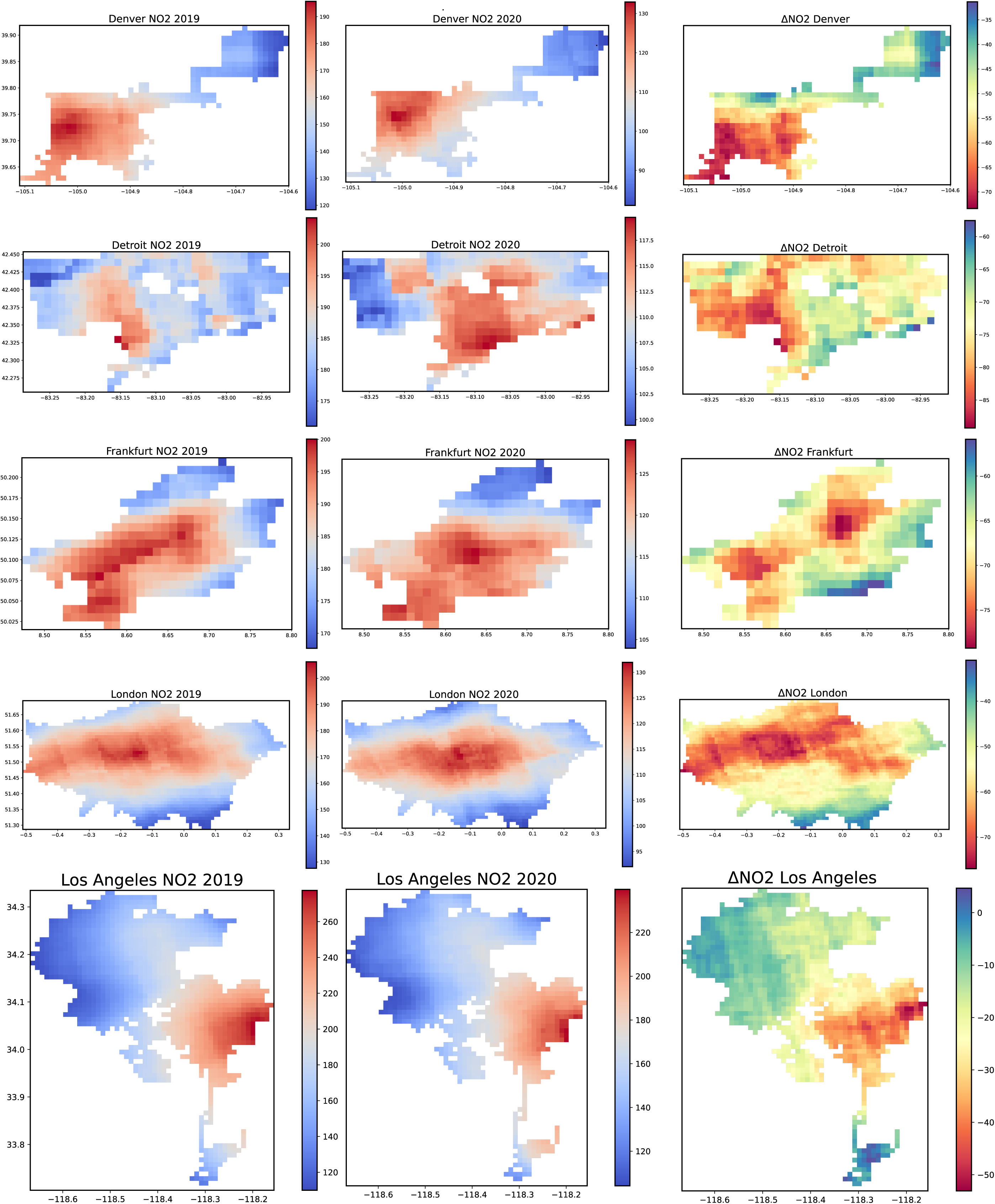
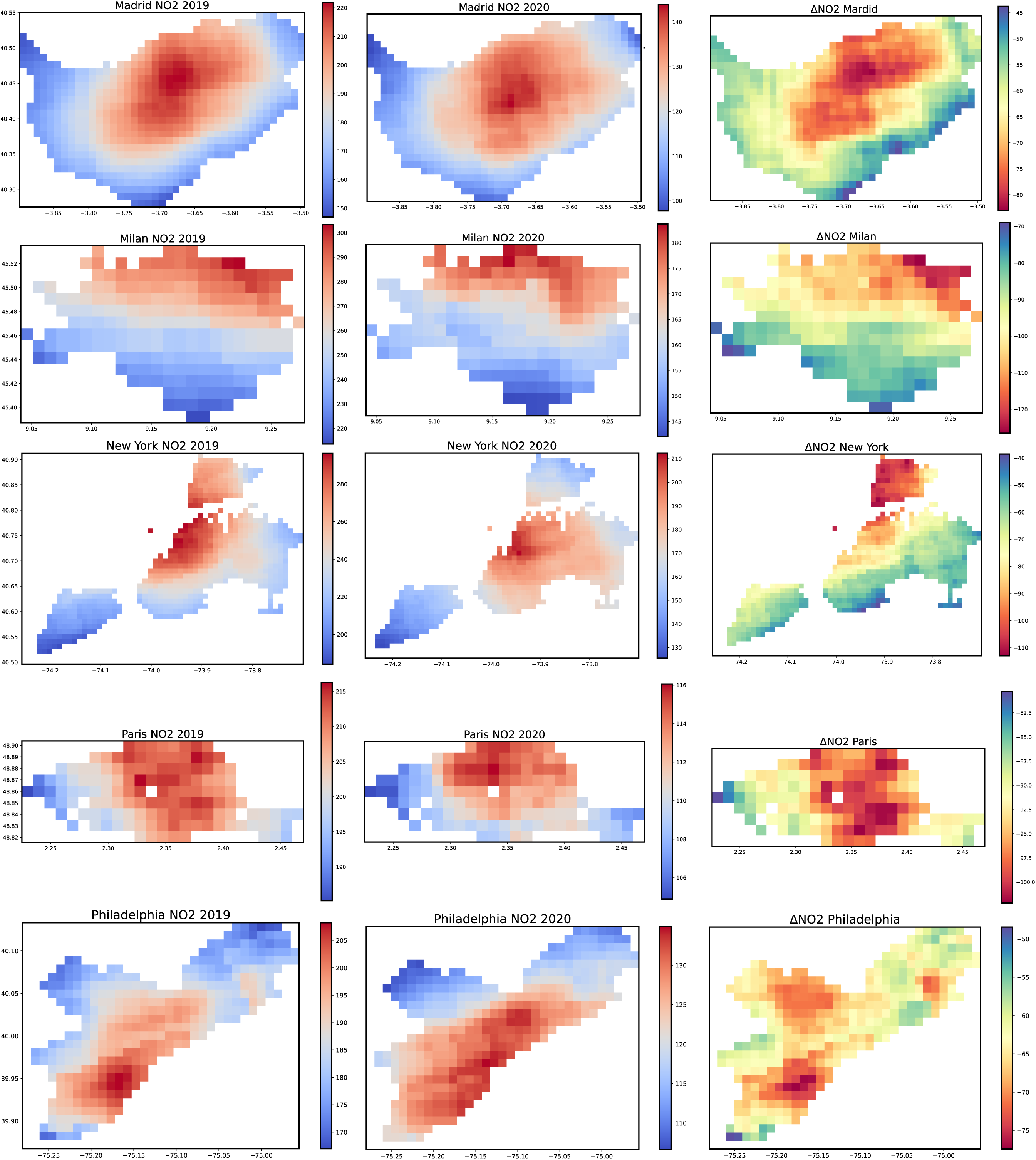
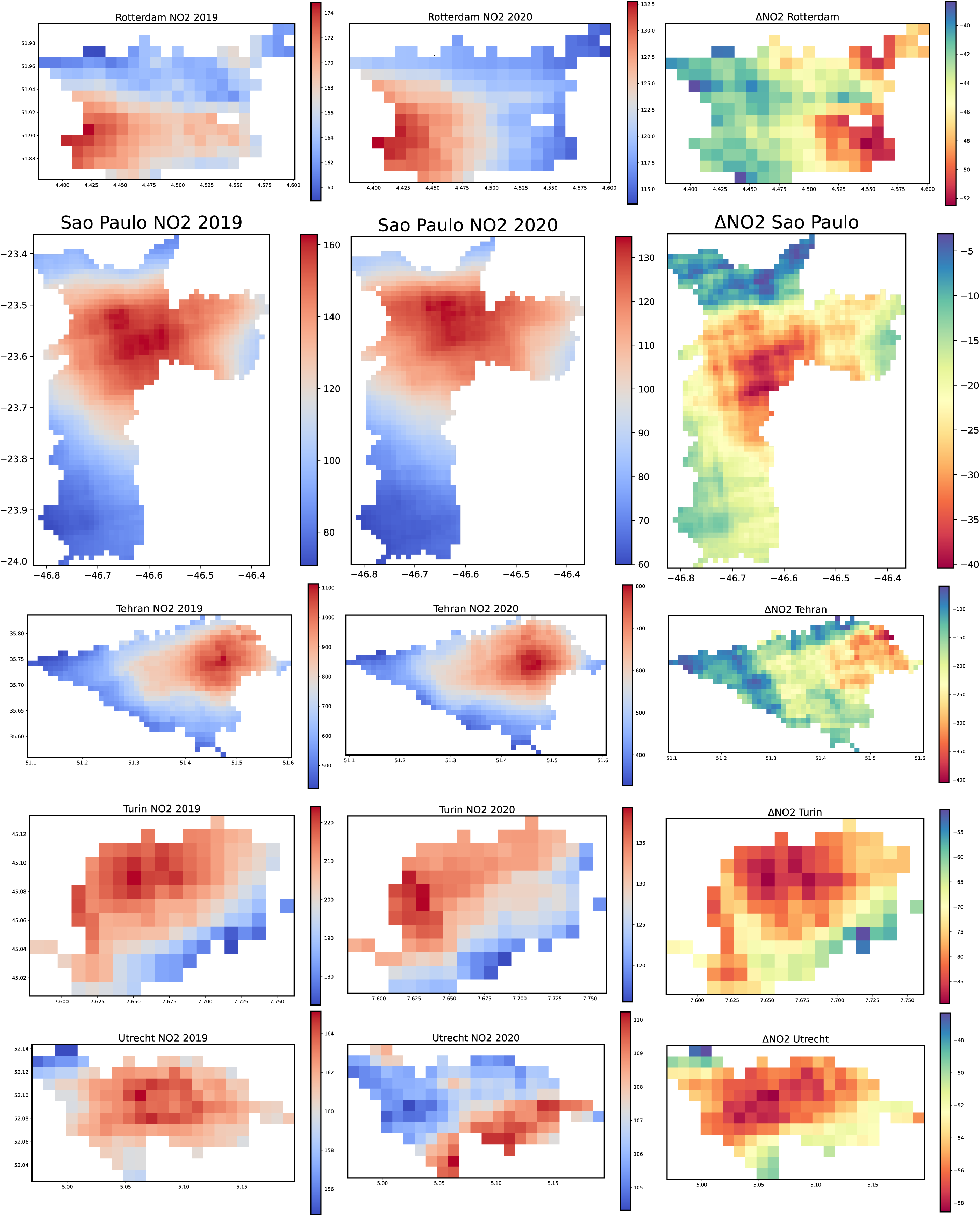
Spatial distribution and changes in NO2 concentration (μmol/m2) derived from Sentinel 5P TROPOMI data.

**Fig. 2.**
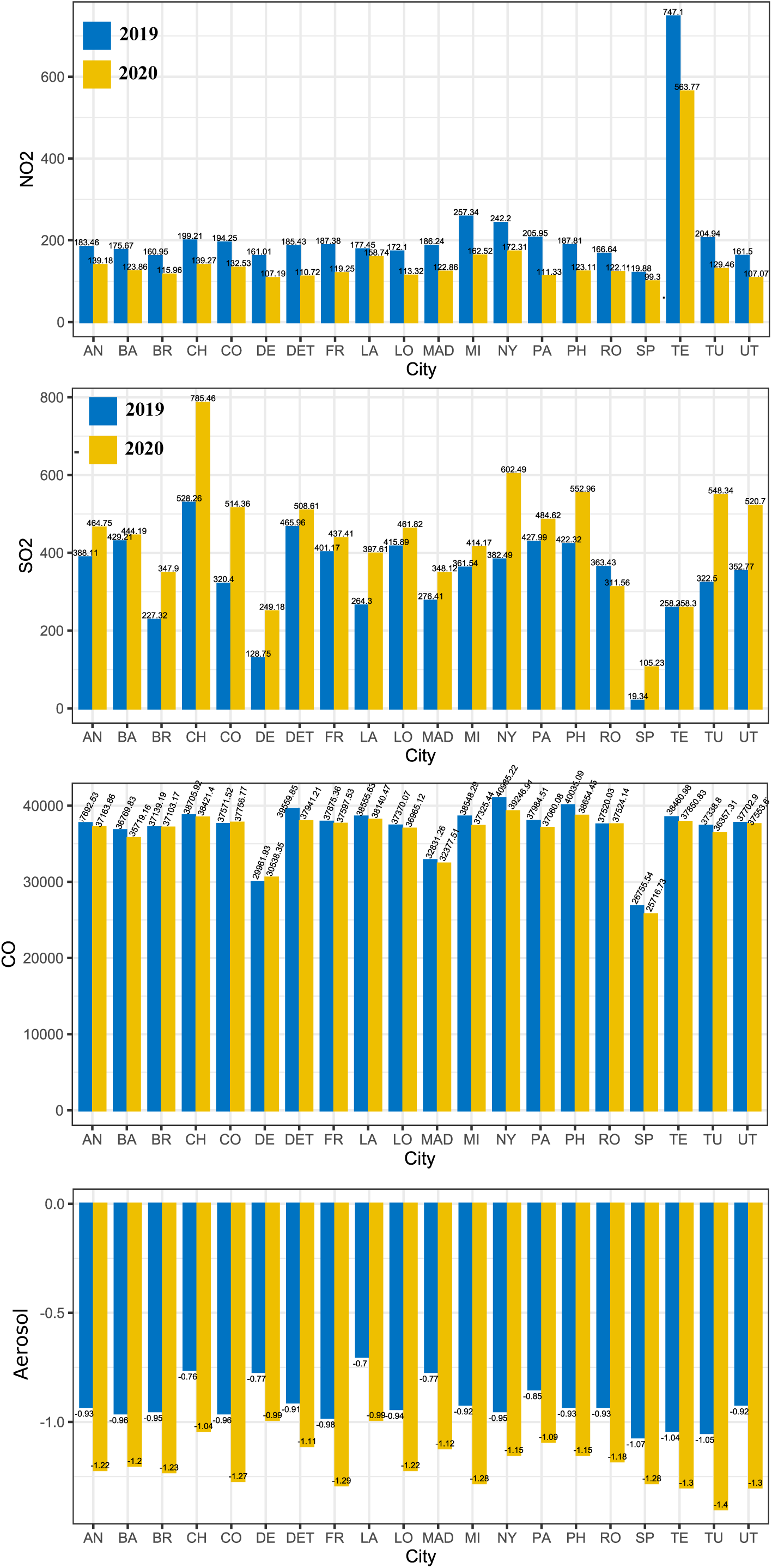
Concentration (μmol/m2) of air pollutants during the study period (1st Feb to 11th May) in 2019 and 2020.

### 3.2 Temporal changes in air pollution due to lockdown

**Fig. 2** and **Table. 1** shows the average NO_2_, SO_2_, CO, and aerosol concentration from 1^st^ Feb to 11^th^ May in 2019 and 2020. Among the 20 cities, the average NO_2_ concentration was found highest in Tehran (747.1µmol in 2019 and 563.77µmol in 2020), followed by Milan (257.34µmol in 2019 and 162.52µmol in 2020), New York (242.2µmol in 2019 and 172.31µmol in 2020), Paris (205.95µmol in 2019 and 111.33µmol in 2020), Turin (204.94µmol in 2019 and 129.46µmol in 2020), Chicago (199.21µmol in 2019 and 139.27µmol in 2020), Cologne (194.25µmol in 2019 and 132.53µmol in 2020), Philadelphia (187.81µmol in 2019 and 123.11µmol in 2020), etc. Lowest NO_2_ concentration was observed in Sao Paulo (119.88µmol in 2019 and 99.3µmol in 2020), Brussels (160.95µmol in 2019 and 115.96µmol in 2020), Denver (161.01µmol in 2019 and 107.19µmol in 2020), respectively. Among the 20 cities, the SO_2_ concentration was found maximum in Chicago (528.26µmol in 2019 and 785.46µmol in 2020), followed by Detroit (465.96µmol in 2019 and 508.61µmol in 2020), Barcelona (429.21µmol in 2019 and 444.19µmol in 2020), Paris (427.99µmol in 2019 and 484.62µmol in 2020), Philadelphia (422.32µmol in 2019 and 552.96µmol in 2020), London (415.89µmol in 2019 and 461.82µmol in 2020), etc. While the low SO_2_ emission was documented in Sao Paulo (19.34µmol in 2019 and 105.23µmol in 2020), Denver (128.75µmol in 2019 and 249.18µmol in 2020), Brussels (227.32µmol in 2019 and 347.9µmol in 2020), Tehran (258.35µmol in 2019 and 258.3µmol in 2020), Los Angeles (264.3µmol in 2019 and 397.61µmol in 2020) (**Fig. 2 and Table. 1**). The average concentration of CO in different cities is also evaluated and presented in **Fig. 2 and Table. 1**. During the study period, the highest CO concentration is recorded in American cities, i.e., New York, Philadelphia, Detroit, Chicago, Los Angeles, while a comparably low CO concentration is documented for Sao Paulo, Denver, Madrid, Barcelona, and Brussels (**Fig. 2**). Except for a few cities, the concentration of NO_2_, CO, and aerosol has been reduced substantially (**Fig. 3 and Table. 1, Table. 2**). For NO_2_, the highest reduction was detected in Paris (45.94%), followed by Detroit (40.29%), Milan(36.85%), Turin (36.83%), Frankfurt (36.36%), Philadelphia (34.45%), London (34.15%), and Madrid (34.03%), respectively. At the same time, comparably lower reduction of NO_2_ is observed in Los Angeles (10.54%), Sao Paulo (17.17%), Antwerp (24.14%), Tehran (24.54%), and Rotterdam (26.72%), respectively (**Fig. 3 and Table. 2**). For CO, the maximum reduction was recorded for New York (4.24%), followed by Detroit (4.09%), Sao Paulo (3.88%), Philadelphia (3.45%), Milan (3.17%), Barcelona (2.86%), respectively. At the same time, a positive (increase) changes in CO were observed in Denver (1.92%), Cologne (0.49%), and Rotterdam (0.01%) (**Fig. 3** and **Table. 2**). The temporal variability of NO_2_, SO_2_, CO, and aerosol concentration is shown in **Fig. 4, Fig. 5, Fig. S4, Fig. S5, Fig. S6, Fig. S7, Fig. S8**. Both median and interquartile range (IQR) values in **Fig. 4** and **Fig. 5** suggest that NO_2_ concentration was decreased substantially. A similar declining pattern is observed for CO for all the 20 cities considered in this study (**Fig. S4, Fig. S5**). However, for SO_2_, an incremental trend was observed for most of the cities (**Fig. S6**).

**Fig. 3.**
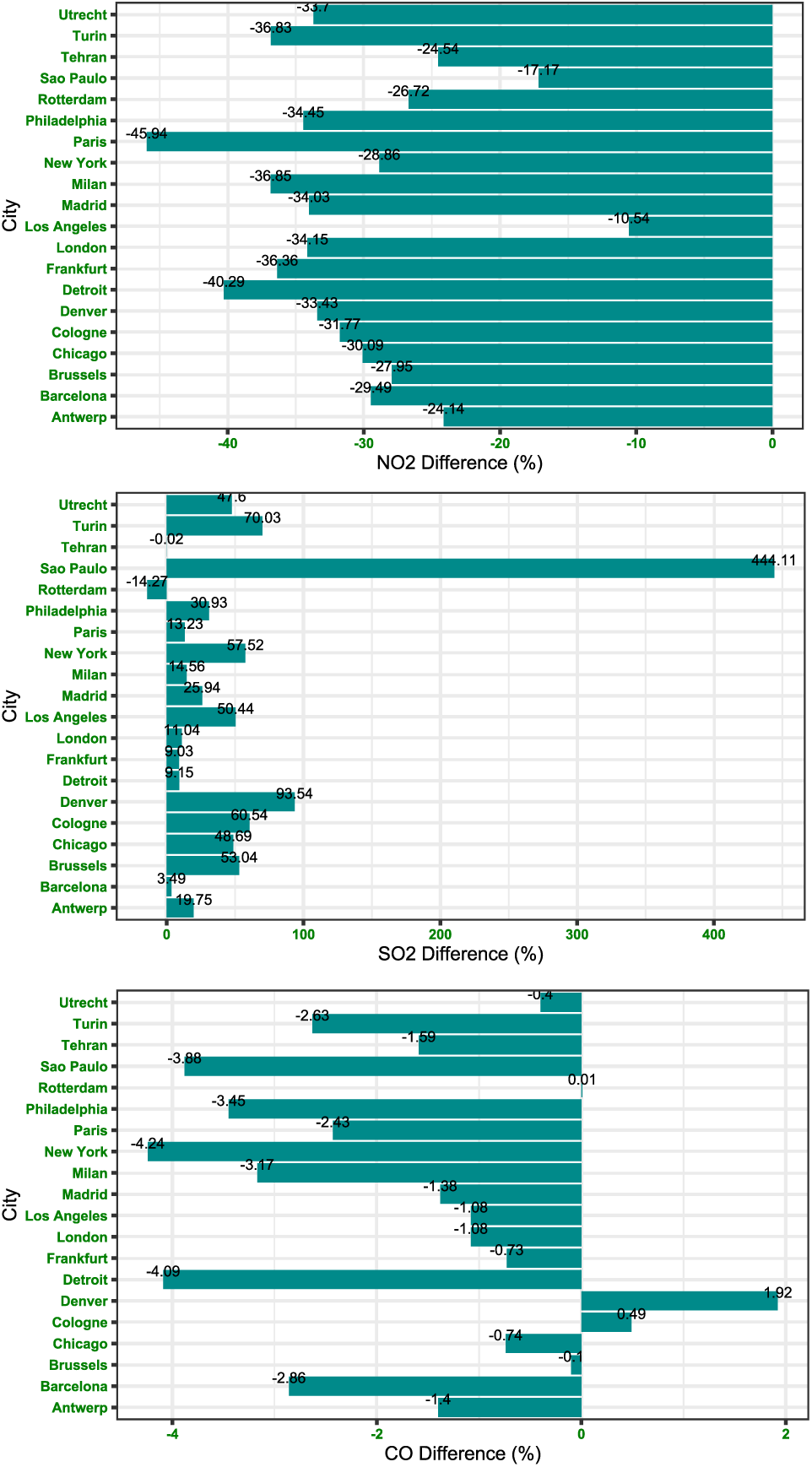
Changes (%) in air pollution during the study period.

**Fig. 4.**
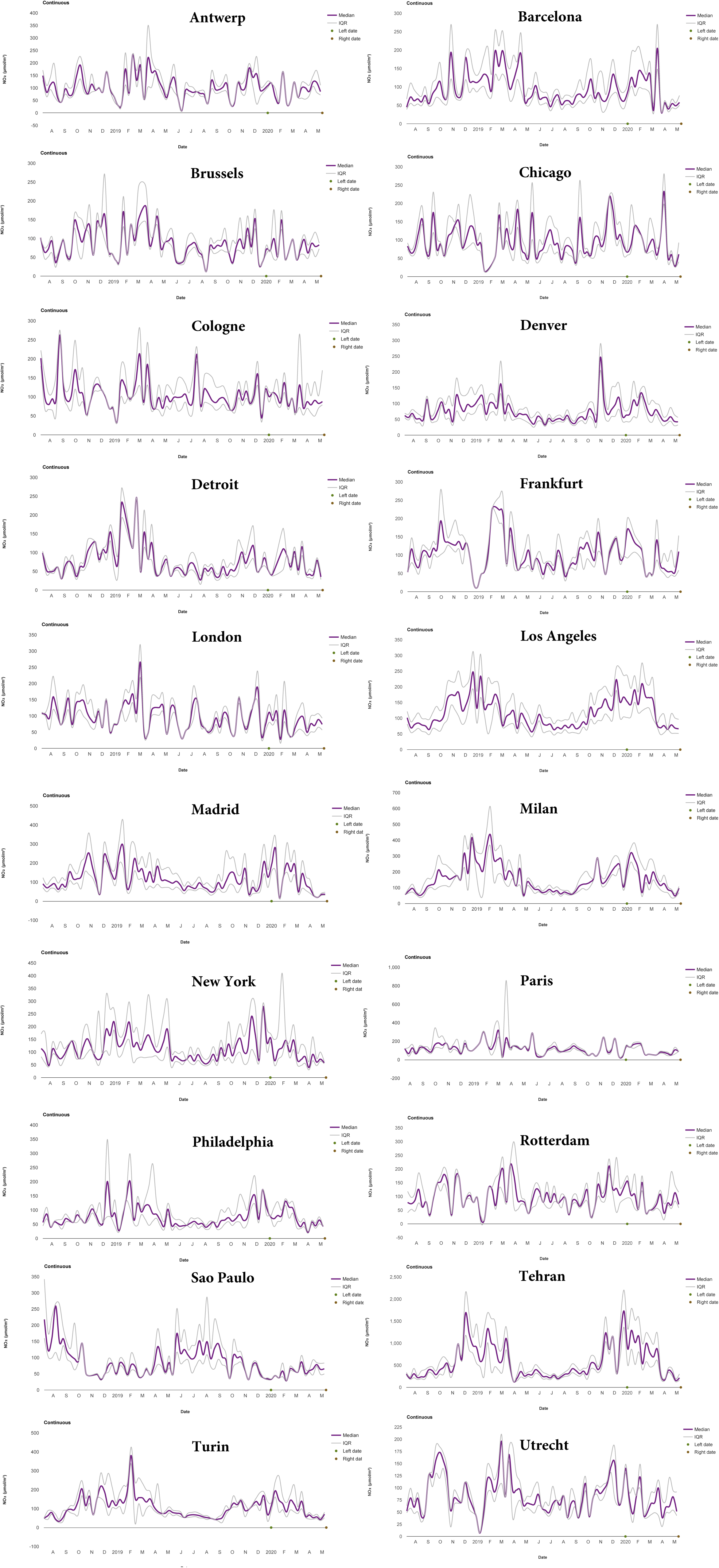
Monthly variation of NO_2_ (μmol m^−2^) concentration in the selected cities from August 2018 to May 2020 derived from Sentinel 5P TROPOMI observation.

**Fig. 5.**
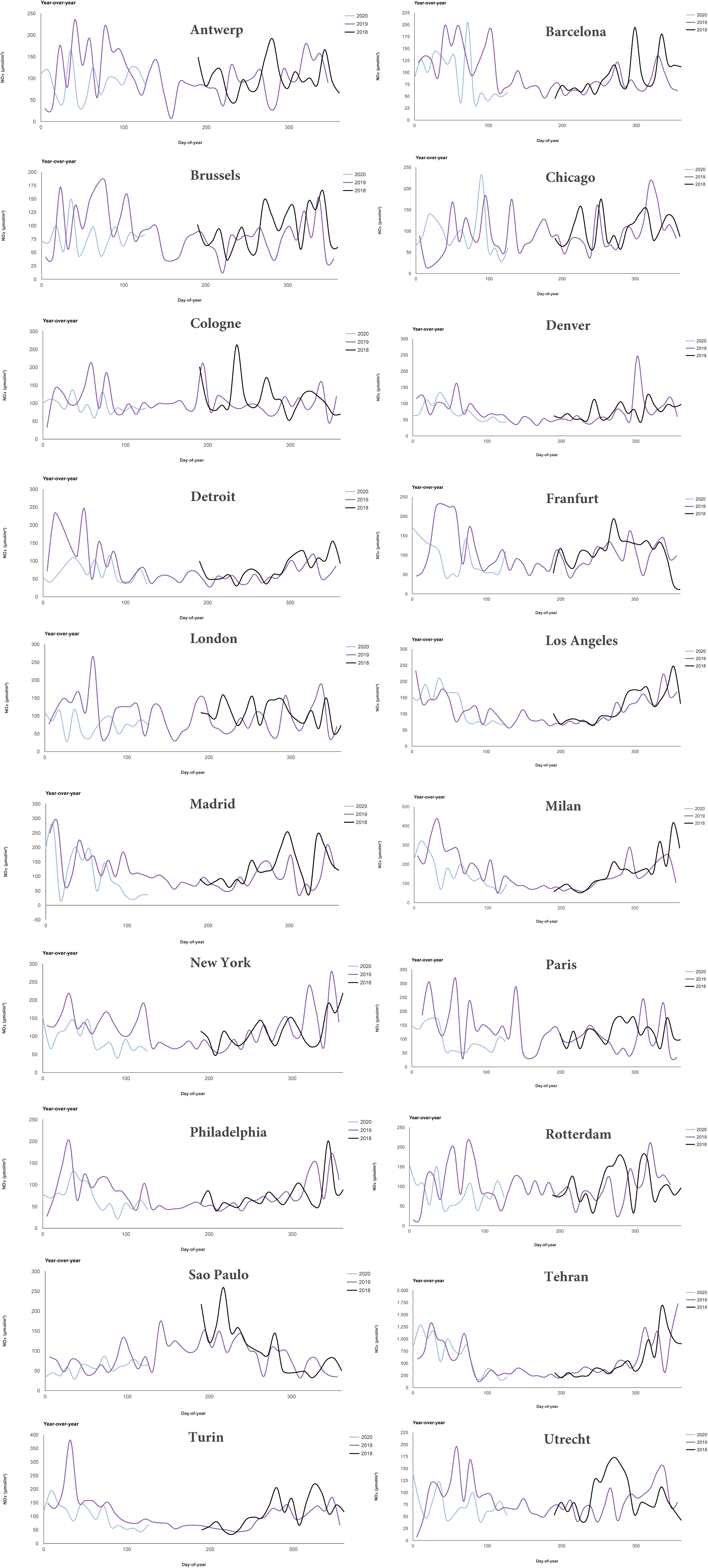
Yearly variation of NO_2_ (μmol m^−2^) concentration in the selected cities in 2018, 2019, and 2020 derived from Sentinel 5P TROPOMI observation.

**Table. 1.**
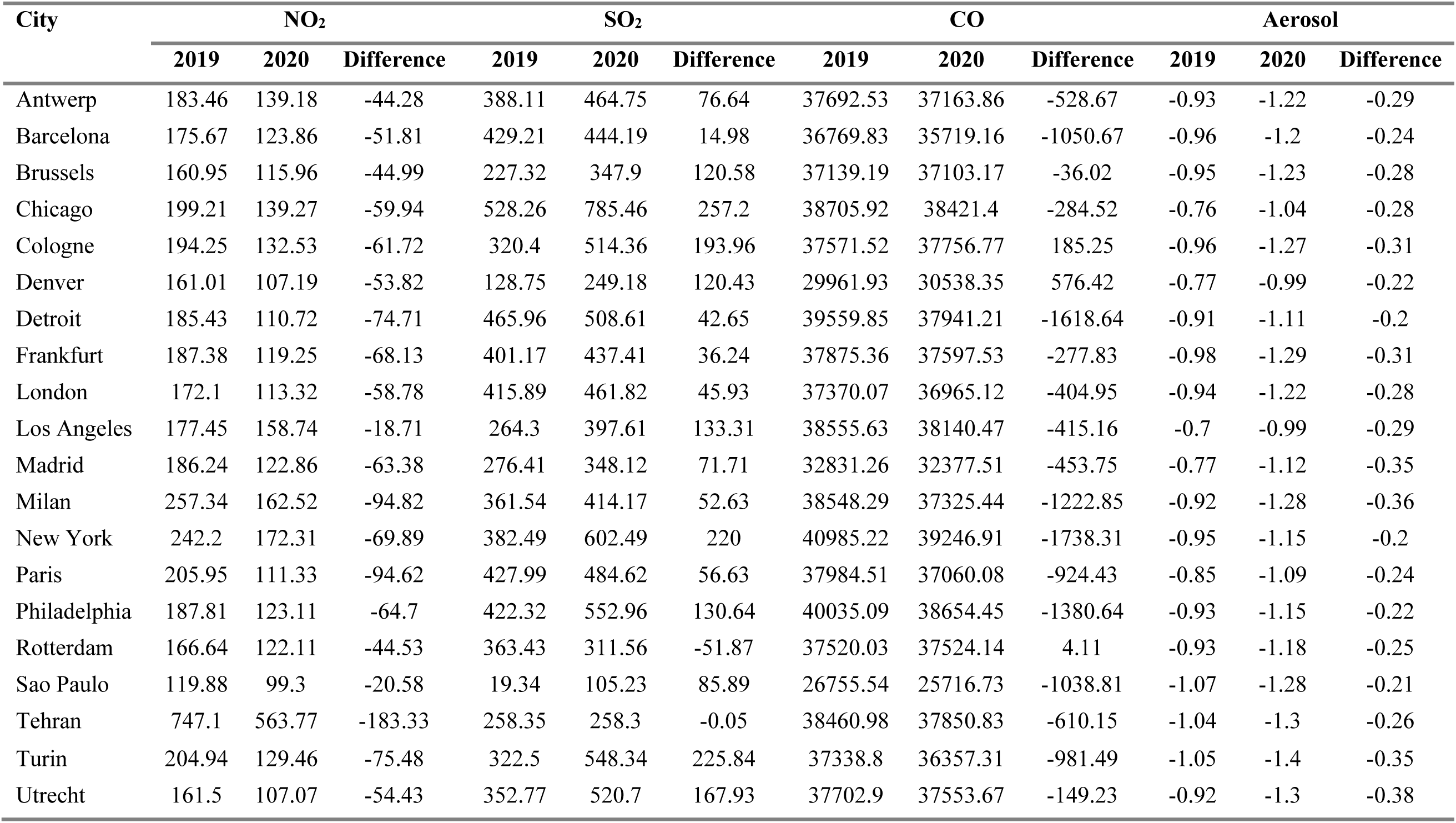
Summary statistics of mean NO_2_, SO_2_, CO, and Aerosol concentration during 2019 and 2020 (Feb to May).

**Table. 2.** Concentration (ton) of different air pollutants in 2019 and 2020 derived from Sentinel TROPOMI satellite data.

| City | NO<sub>2</sub> |  |  | SO<sub>2</sub> |  |  | CO |  |  |
| --- | --- | --- | --- | --- | --- | --- | --- | --- | --- |
|  | 2019 | 2020 | Difference | 2019 | 2020 | Difference | 2019 | 2020 | Difference |
|  |  |  | (%) |  |  | (%) |  |  | (%) |
| Antwerp | 1.73 | 1.31 | -24.14 | 5.08 | 6.09 | 19.75 | 215.83 | 212.80 | -1.40 |
| Barcelona | 0.82 | 0.58 | -29.49 | 2.80 | 2.90 | 3.49 | 104.91 | 101.91 | -2.86 |
| Brussels | 1.20 | 0.86 | -27.95 | 2.35 | 3.60 | 53.04 | 167.84 | 167.68 | -0.10 |
| Chicago | 5.55 | 3.88 | -30.09 | 20.51 | 30.50 | 48.69 | 656.87 | 652.04 | -0.74 |
| Cologne | 3.62 | 2.47 | -31.77 | 8.32 | 13.35 | 60.54 | 426.27 | 428.37 | 0.49 |
| Denver | 2.97 | 1.98 | -33.43 | 3.31 | 6.40 | 93.54 | 336.58 | 343.06 | 1.92 |
| Detroit | 3.16 | 1.89 | -40.29 | 11.05 | 12.06 | 9.15 | 409.95 | 393.18 | -4.09 |
| Frankfurt | 2.14 | 1.36 | -36.36 | 6.38 | 6.96 | 9.03 | 263.32 | 261.39 | -0.73 |
| London | 12.45 | 8.20 | -34.15 | 41.88 | 46.51 | 11.04 | 1644.88 | 1627.06 | -1.08 |
| Los Angeles | 10.63 | 9.51 | -10.54 | 22.05 | 33.17 | 50.44 | 1405.58 | 1390.45 | -1.08 |
| Madrid | 5.18 | 3.42 | -34.03 | 10.70 | 13.48 | 25.94 | 555.52 | 547.84 | -1.38 |
| Milan | 2.15 | 1.36 | -36.85 | 4.21 | 4.82 | 14.56 | 196.23 | 190.00 | -3.17 |
| New York | 8.73 | 6.21 | -28.86 | 19.21 | 30.25 | 57.52 | 899.48 | 861.33 | -4.24 |
| Paris | 1.00 | 0.54 | -45.94 | 2.89 | 3.27 | 13.23 | 112.10 | 109.37 | -2.43 |
| Philadelphia | 3.17 | 2.08 | -34.45 | 9.93 | 13.00 | 30.93 | 411.40 | 397.21 | -3.45 |
| Rotterdam | 2.50 | 1.83 | -26.72 | 7.59 | 6.50 | -14.27 | 342.27 | 342.31 | 0.01 |
| Sao Paulo | 8.39 | 6.95 | -17.17 | 1.88 | 10.25 | 444.11 | 1139.46 | 1095.22 | -3.88 |
| Tehran | 25.09 | 18.93 | -24.54 | 12.08 | 12.08 | -0.02 | 786.14 | 773.67 | -1.59 |
| Turin | 1.23 | 0.78 | -36.83 | 2.69 | 4.57 | 70.03 | 136.12 | 132.54 | -2.63 |
| Utrecht | 0.74 | 0.49 | -33.70 | 2.24 | 3.31 | 47.60 | 104.73 | 104.32 | -0.40 |

Using the ground monitored data, the daily air quality index (AQI), and a cumulative number of good AQI days for the six American cities was computed and presented in **Fig. 6**. The ground monitored data for these six cities have been considered only for time series assessment and subsequent interpretation. In all cases, it has been found that AQI is reduced significantly due to lockdown led reduction in human mobility and traffic emission. In the left panel, the grey color indicates the five years average AQI and light blue shade demonstrating the average AQI range in the last 20 years. Based on the AQI ranges, four AQI classes were characterised, such as good, moderate, unhealthy for sensitive groups, and unhealthy (**Fig. 6**). A comparably higher cumulative number of good AQI days is recorded during the lockdown period for all five cities, except Chicago (**Fig. 6**). Using the EPA AQI interactive plot function application, the daily AQI of the US cities were analysed and presented in **Fig. 7, Fig. S9, Fig. S10**. The daily NO_2_ and SO_2_ AQI suggest that all the cities are benefitted by having good quality air due to anthropogenic pollution switch-off and restricted human mobility that collectively improved the air quality ecosystem services in these cities. The multi-year daily time series plot (**Fig. 8, Fig. 9, Fig. 10**) is also indicating the improving status of air quality in the US cities due to the reduced level of traffic emission. Six distinct color grade is used to demonstrate the AQI categories. Six different AQI classes, i.e., good, moderate, unhealthy for sensitive groups, unhealthy, very unhealthy, and hazardous, etc. are also defined to evaluate the time series AQI status of these cities during the pre-COVID (2000 – 2019) and lockdown (Jan to May 2020) period. **Fig. 8** shows that in all cities, the NO_2_ AQI status is mostly good during the lockdown period compared to the long-term average AQI in these cities. **Fig. 9** shows the PM_2.5_ AQI status, which also found improving during the lockdown period. The higher proportions of good AQI values in all the cities are suggesting improving air quality (PM_2.5_) status in Chicago, Denver, Detroit, Los Angeles, New York, and Philadelphia. The multi-year time series plot was prepared after combining all the pollutants that suggest that the air quality is improved substantially, which is supported by the lower AQI recorded during the lockdown period compared to the long-term AQI recorded in these cities. Among the six cities, the hazardous to very unhealthy air quality is common in Los Angeles, compared to the other five US cities considered in this study.

**Fig. 6.**
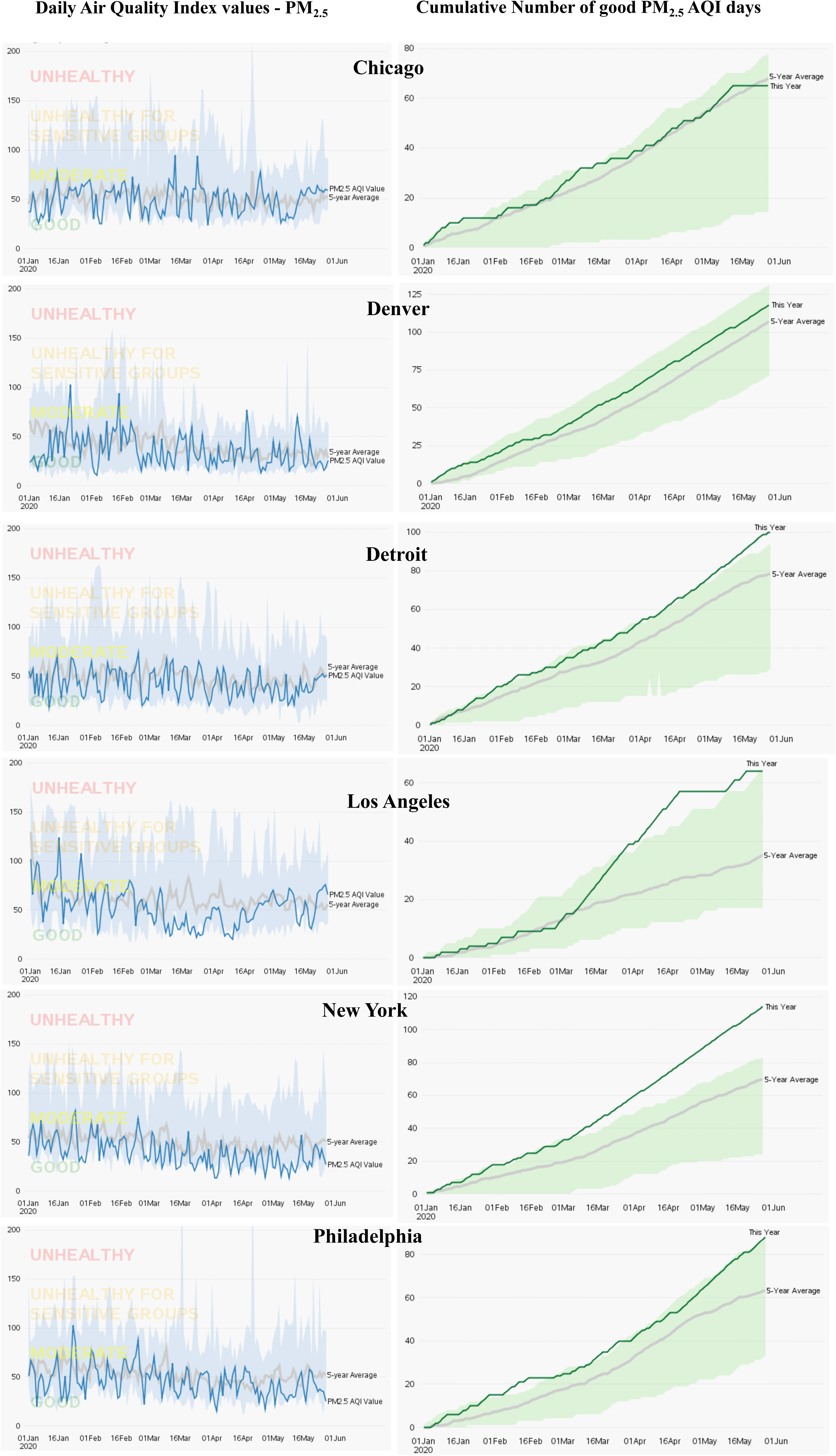
Shows the ground data based PM_2.5_ air quality index values for the selected cities. Figures in left panel shows the 20 years (2000 – 2019) air quality index values, 5 years average (2015 – 2019) and most recent PM_2.5_ air quality index values of the selected cities. The maps on the right panel shows recent (green color) and 5 years average (gray color) cumulative number of good PM_2.5_ air quality index days for the selected cities. Maps in both panel are indicating the improving status of air quality in the selected cities.

**Fig. 7.**
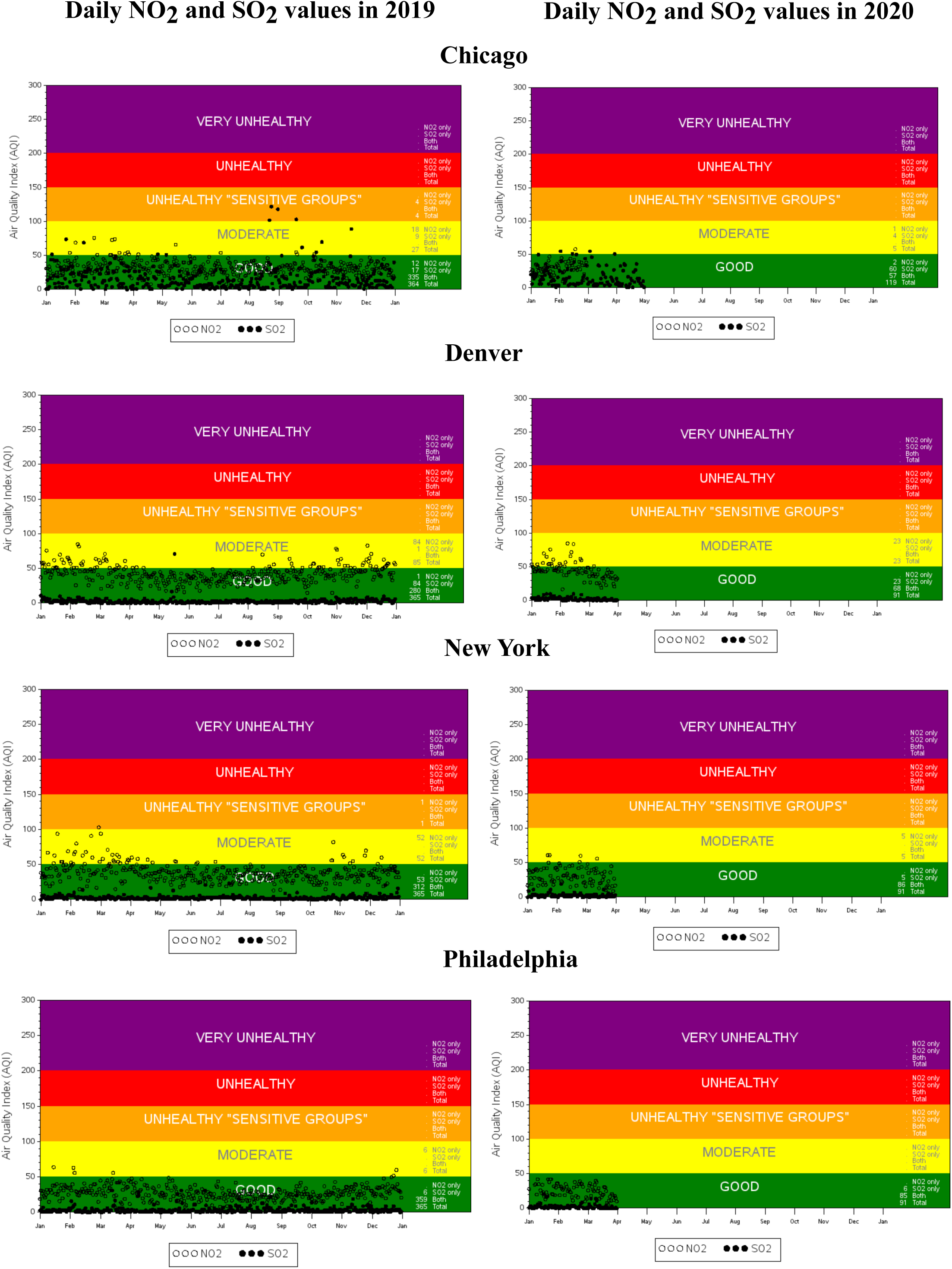
Shows the ground monitored air quality index values of NO_2_ and SO_2_ in 2019 and 2020 in the selected cities. In most cases, air quality has been improved substantially during the lock down period.

**Fig. 8.**
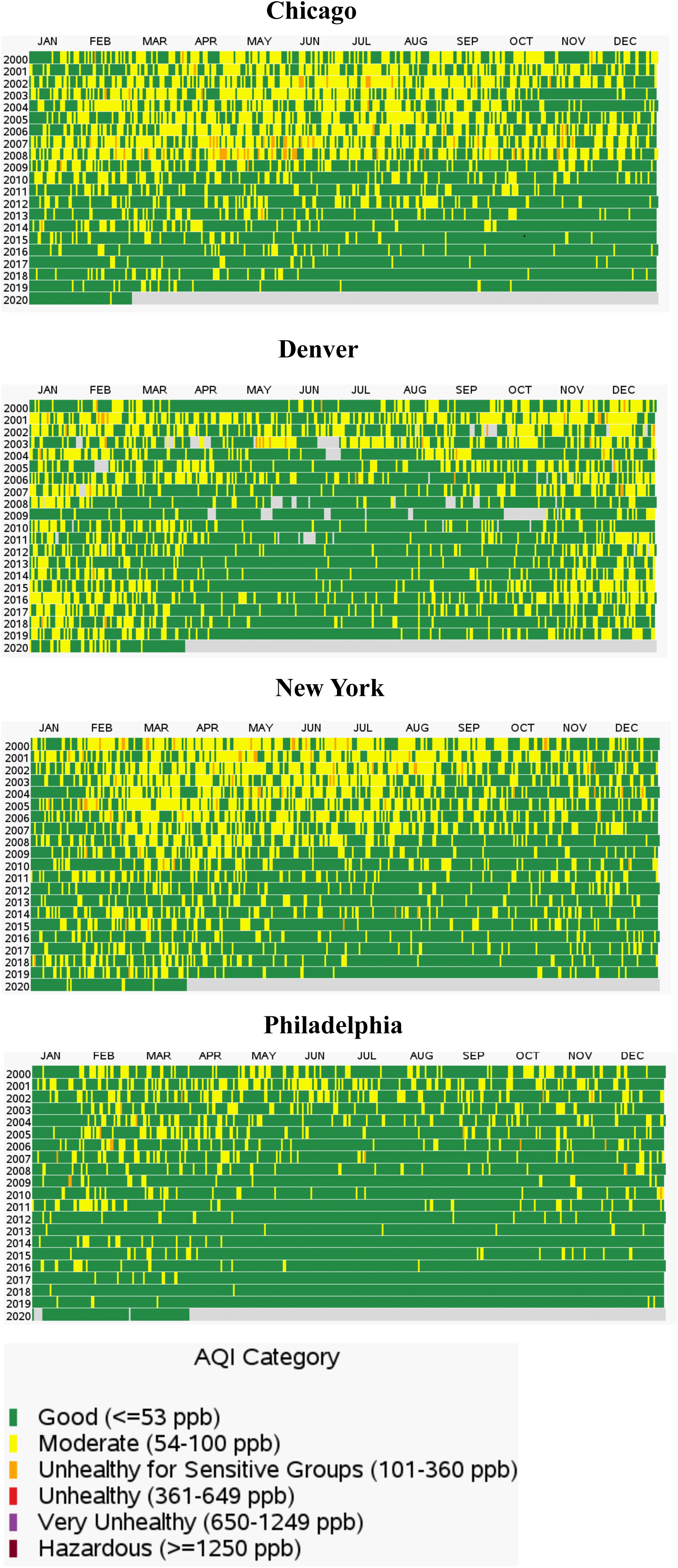
Multi-year daily time series plot shows the variation of air quality status (NO_2_) from 2000 to 2020. Due to lock down and associated reduction of air pollution, air quality status is improved in all the selected cities in USA.

**Fig. 9.**
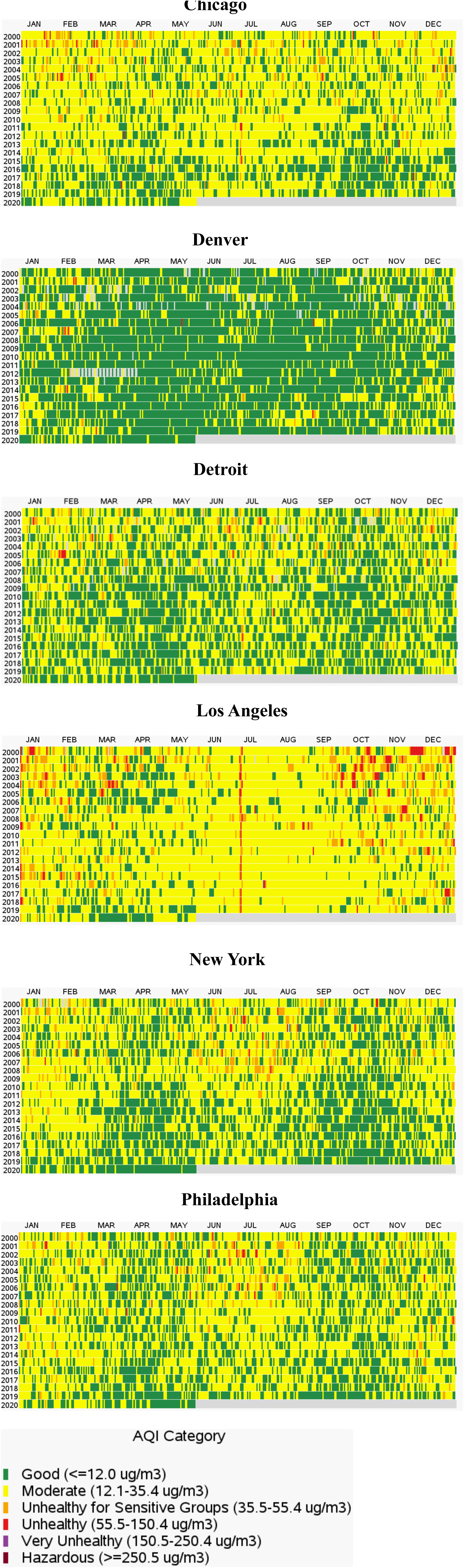
Multi-year daily time series plot shows the variation of air quality status (PM_2.5_) from 2000 to 2020. Due to lock down and associated reduction of air pollution, air quality status is improved in all the selected cities in USA.

**Fig. 10.**
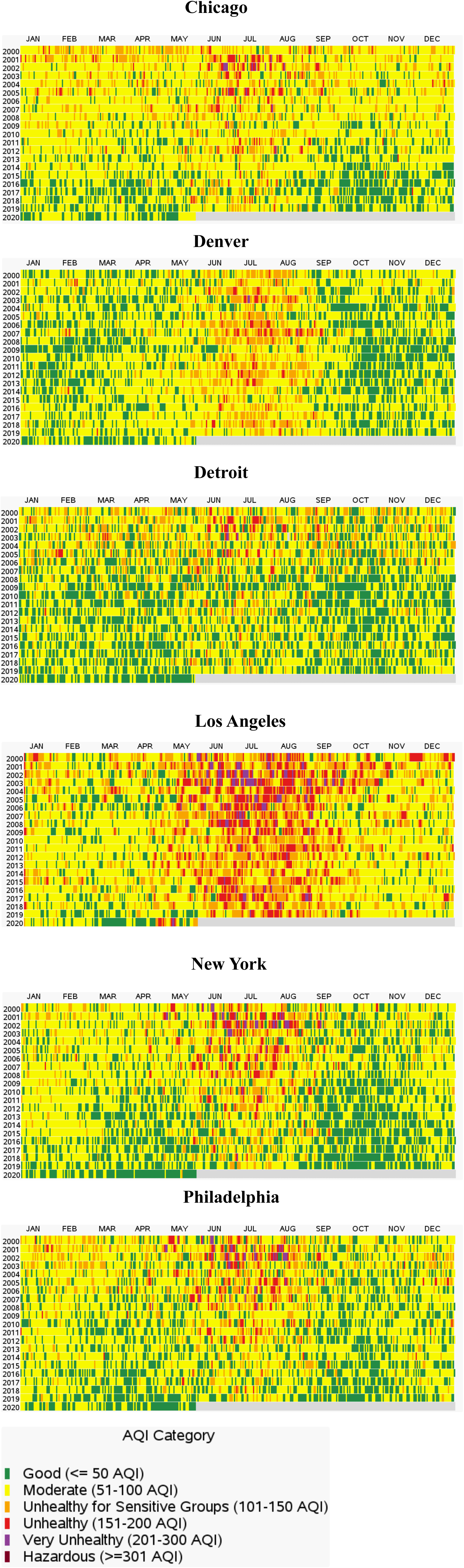
Multi-year daily time series plot shows the variation of air quality status (after considered all pollutants) from 2000 to 2020. Due to lock down and associated reduction of air pollution, air quality status is improved in all the selected cities in USA.

### 3.3 Human mobility and its paramount effect in lowering the pollution levels

Using both Google and Apple human mobility information, the effect of lockdown and its striking impact on human outdoor activities is measured and presented in **Fig. 11, Fig. S12, Fig. S13, Fig. S14, Fig. S15**. The driving and transit mobility was calculated using the Apple mobility data. Mobility on January 13 was taken as a baseline, and further changes in human mobility during the lockdown period was calculated from the baseline mobility. The driving counts reduced most significantly in Paris, followed by Madrid, London, Antwerp, and Brussels (**Fig. 11**). Whereas, such changes were comparably lower in Chicago, Cologne, Denver, Los Angeles, New York (**Fig. 11**). Transit counts also reduced significantly in Paris, followed by Utrecht, Sao Paulo, New York, Milan, Chicago, Antwerp, and Brussels (**Fig. 11**). Using the Google human mobility records, the changes in different mobility such as retail and recreation, grocery and pharmacy stores, transit, parks and outdoor, workplace visitor, and time spent at home were measured. Transport related mobilities were reduced most significantly in the Latin American countries, followed by a few Middle East and Southeast Asian countries, and American countries (**Fig. S13**). Parks and outdoor activities were found to be reduced maximum in the Latin American countries and South Asian countries. At the same time, outdoor activities are seen to be increased in a few European countries as well (**Fig. S13**). The highest reduction in retail and recreation is found in India, Turkey, UK, and few Latin American countries due to lockdown and associated restrictive measures. (**Fig. S14**). Considering grocery and pharmacy-related mobilities, the highest reduction is being observed in the Latin American countries and a few European countries. Whereas grocery related mobility was found to be increased in the USA, few African and European countries (**Fig. S14**). Workplace related mobility is reduced significantly in Peru, Bolivia, India, Spain, Turkey, Saudi Arabia, USA, and Canada (**Fig. S15**). While such changes were positive in a few African countries (Mali, Niger, Mozambique, Zambia), Venezuela, and a few island countries (**Fig. S15**). Finally, using the Google real-time mobility information, another mobility component, i.e., time spent at home, was calculated (**Fig. S15**). As expected, due to lockdown and mandatory restrictive measures on human activities, people tend to spend more time at home, which also suggests that at most of the countries have taken timely decisions to control the pandemic. Except for a few European countries, peoples around the world limited their outdoor activities, which is supported by the results shown in **Fig. S15**.

**Fig. 11.**
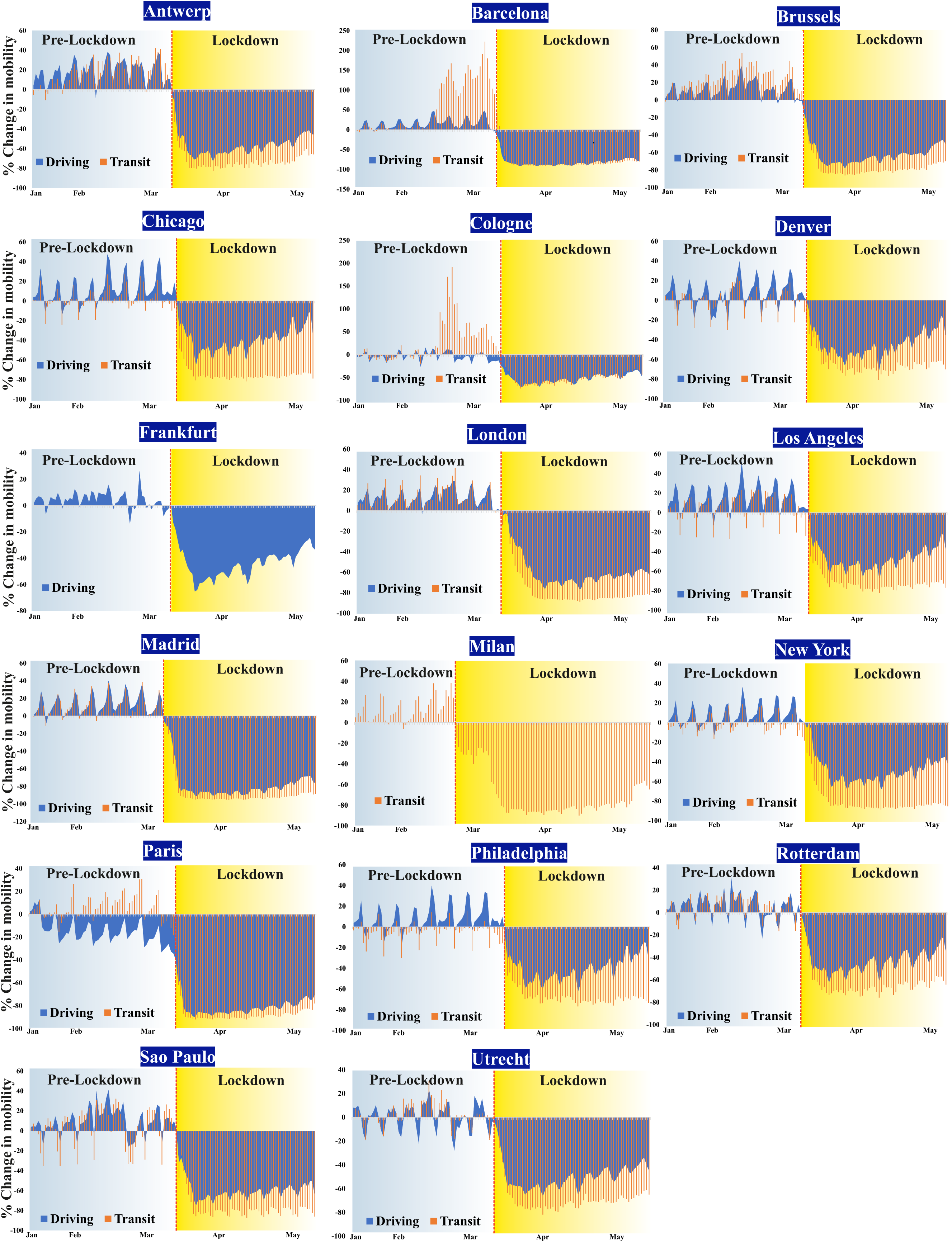
Changes in mobility due to lock down led restriction in the selected cities.

### 3.4 Improving the status of air quality ecosystem services

Using both public health and externality valuation approaches, the positive association between lockdown led the reduction of anthropogenic emissions, and air quality ecosystem services are analysed and presented in **Table. 3, Table. 4, Table. 5, Table. S5**. Before economic valuation, the original externality values for different air pollutants were adjusted using the latest price inflation conversion factor (**Table. 3)**. These adjusted value coefficients were later used to calculate the economic value of the air quality ecosystem services for the 20 cities across the world. For the public health valuation method, the estimated economic burden and economic benefits were also adjusted for eliminating the influence of price inflation in the valuation. Overall, the per-unit EV was calculated maximum for Sao Paulo (49716 $), New York (49453 $), Tehran (43624 $), London (38930 $), Detroit (22588 $), Los Angeles (20242 $), Philadelphia (19190 $), Madrid (16413 $), Chicago (13222 $), Milan (10035 $), Frankfurt (5854 $), Turin (5749 $), Antwerp (5039 $), Paris (4971 $), Barcelona (4117 $), Cologne (3914 $), Rotterdam (3400 $), Brussels (1876 $), and Utrecht (1675 $). At the same time, the economic burden (both NO_2_ and CO emission is found higher than the previous year, 2019) due to NO_2_ and CO emission was calculated for Denver (−1077 $) (**Table. 4**).

**Table. 3.** Per unit ecosystem service equivalent value of different pollutants.

| Pollutants | Min | Median | Mean | Max |
| --- | --- | --- | --- | --- |
| CO | \$1.84 | \$956.17 | \$956.17 | \$1,930.72 |
| NO<sub>x</sub> | \$404.53 | \$1,949.11 | \$5,148.58 | \$17,468.41 |
| SO<sub>2</sub> | \$1,415.86 | \$3,309.80 | \$3,677.56 | \$8,642.27 |
| PM<sub>10</sub> | \$1,746.84 | \$5,148.58 | \$7,906.76 | \$29,788.24 |

**Table. 4.** Economic benefits due to the reduction of anthropogenic emission estimated for different cities estimated using median externality valuation method.

| City | NO<sub>2</sub> | CO | Overall |
| --- | --- | --- | --- |
|  | ESV (USD) | ESV (USD) | ESV (USD) |
| Antwerp | 2145 | 2894 | 5039 |
| Barcelona | 1251 | 2866 | 4117 |
| Brussels | 1720 | 156 | 1876 |
| Chicago | 8605 | 4617 | 13222 |
| Cologne | 5924 | -2010 | 3914 |
| Denver | 5114 | -6191 | -1077 |
| Detroit | 6549 | 16038 | 22588 |
| Frankfurt | 4007 | 1847 | 5854 |
| London | 21887 | 17043 | 38930 |
| Los Angeles | 5770 | 14472 | 20242 |
| Madrid | 9072 | 7341 | 16413 |
| Milan | 4083 | 5952 | 10035 |
| New York | 12975 | 36478 | 49453 |
| Paris | 2362 | 2609 | 4971 |
| Philadelphia | 5624 | 13566 | 19190 |
| Rotterdam | 3436 | -36 | 3401 |
| Sao Paulo | 7414 | 42302 | 49716 |
| Tehran | 31700 | 11925 | 43624 |
| Turin | 2328 | 3421 | 5749 |
| Utrecht | 1279 | 396 | 1675 |

**Table. 5.** Summary estimates of economic benefits (Million US$) derived from health burden approach. EB = Economic Burden (Million US$)

| City | EB 2019 | EB 2020 | Economic Benefit |
| --- | --- | --- | --- |
| Antwerp | 67 | 51 | 16 |
| Barcelona | 138 | 97 | 41 |
| Brussels | 31 | 22 | 9 |
| Chicago | 456 | 320 | 137 |
| Cologne | 213 | 145 | 67 |
| Denver | 89 | 59 | 30 |
| Detroit | 106 | 64 | 43 |
| Frankfurt | 144 | 92 | 52 |
| London | 1102 | 727 | 375 |
| Los Angeles | 634 | 568 | 67 |
| Madrid | 267 | 176 | 90 |
| Milan | 211 | 134 | 78 |
| New York | 1744 | 1243 | 501 |
| Paris | 270 | 146 | 124 |
| Philadelphia | 258 | 169 | 89 |
| Rotterdam | 35 | 26 | 9 |
| Sao Paulo | 234 | 194 | 40 |
| Tehran | 152 | 115 | 37 |
| Turin | 123 | 78 | 45 |
| Utrecht | 41 | 27 | 14 |

The population-weighted average concentration (PWAC, µmol m^−2^) was estimated for each city and presented in **Fig. 12**. The highest PWAC values (in 2019) were estimated for Tehran (512), followed by Milan (183), New York (139), Chicago (139), Turin (139), Philadelphia (134), Los Angeles (133), Madrid (133), Paris (133), Detroit (127), Cologne (126), London (125), Frankfurt (122), respectively. Using the public health burden valuation approach, the highest economic values (derived from public health burden valuation approach and estimated for 101 days) was estimated for New York (501M $), followed by London (375 M $), Chicago (137M $), Paris (124M $), Madrid (90M $), Philadelphia (89M $), Milan (78 M $), Cologne (67M $), Los Angeles (67M $), Frankfurt (52M $), Turin (45M $), Detroit (43 M $), Barcelona (41M $), Sao Paulo (40M $), Tehran (37M $), Denver (30M $), Antwerp (16 M $), Utrecht (14M $), Brussels (9M $), and Rotterdam (9M $), respectively (**Table. 5**). It is also evident from the economic valuation that due to the temporary reduction of air pollution levels, the economic cost attributed to air pollution led health burdens was reduced significantly (**Table. S5**).

**Fig. 12.**
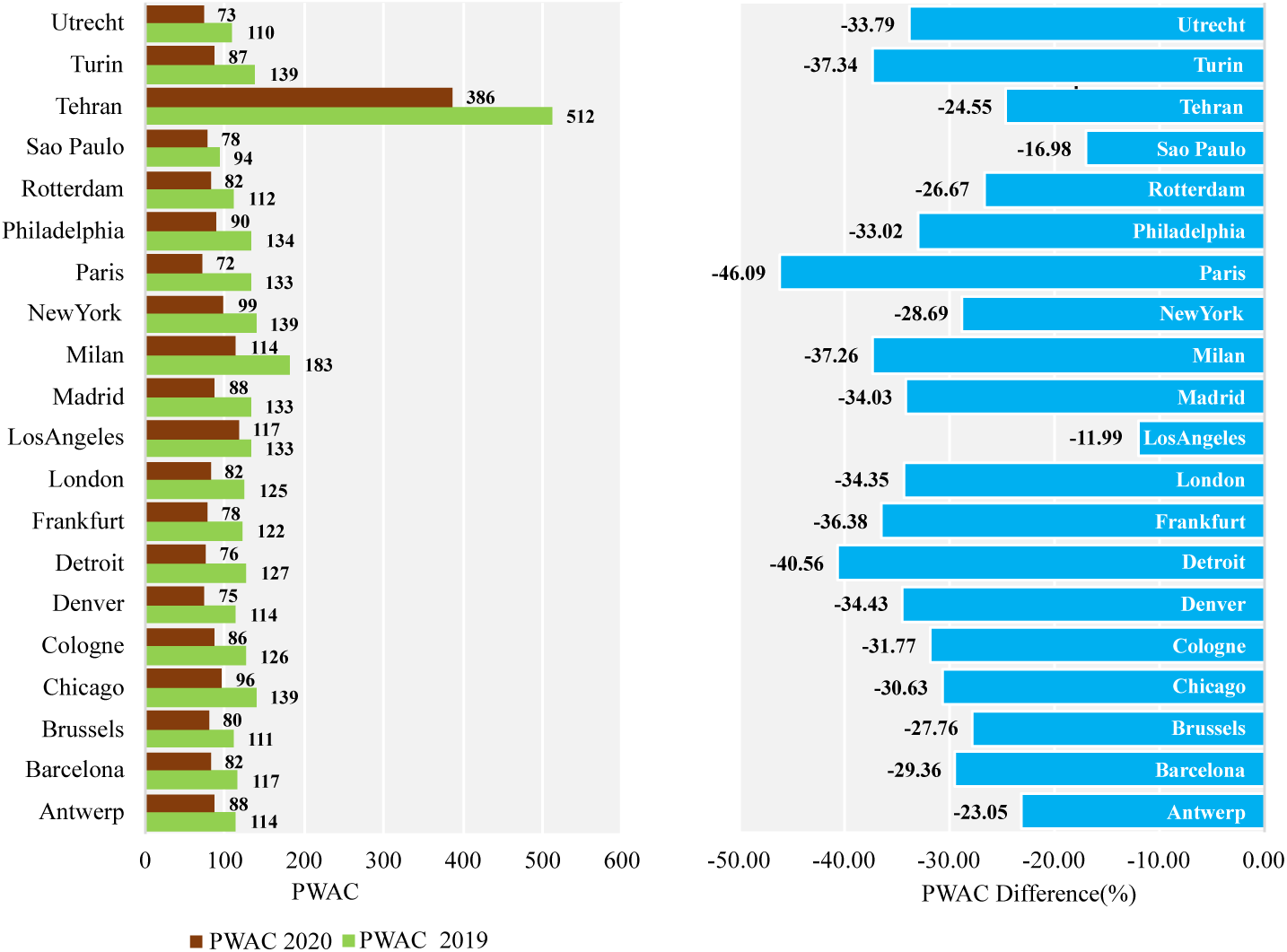
Pollution weighted average concentration of the major cities in 2019 and 2020 (during Feb 1 to May 11).

## 4. Discussion

### 4.1 Relevance of satellite remote sensing in air pollution mapping

Using the ESA Sentinel 5P TROPOMI air real-time pollution data, the spatiotemporal concentration of different air pollutants, i.e., NO_2_, SO_2_, CO, Aerosol, has been evaluated to examine the positive effects of COVID-19 lockdown on air quality across the world. Sentinel 5P satellite mission is one of the finest space-borne applications that provide the crucial key information of air quality, ozone, ultra-violate radiation, and climate monitoring and forecasting (ESA, 2020). TROPOMI widens the application of the satellite air pollution observation and works in line with other global missions, i.e., SCIAMACHY (2002–2012), GOME-2 (since 2007), and OMI (since 2004) (Lorente et al., 2019). This data has been used for many purposes, including air pollution measurement (Zheng et al., 2019; Borsdorff et al., 2018; Shikwambana et al., 2020), epidemiological studies (Chen et al., 2020; Dutheil et al., 2020b; Gautam, 2020; Muhammad et al., 2020; Ogen, 2020; Shehzad et al., 2020); monitoring global volcano (Valade et al., 2019), demographic analysis (Kaplan and Yigit, 2020), evaluating sun-induced chlorophyll fluorescence (SIF) (Guanter et al., 2015), estimation of volcanic sulfur dioxide emission (Theys et al., 2019), etc. In addition, the advent of Google Earth Engine cloud-based suitability in handling the large volume of spatial data facilitates the application of satellite images for timely decision making and offering cost-benefit solutions to many environmental problems. Furthermore, most of the fine to medium scale satellite data products are free and open access in nature (Woodcock et al., 2008). This suggests that transferring ideas from place to place would be easy, which eventually establishes more trust and transparency in applying the scientific findings to solve real-life problems. Evaluating the reliability of remote sensing data is always a matter of concern. Since this study has evaluated the air pollution in cities, which itself is very sensitive in nature, proper and careful evaluation is required to verify the accuracy of satellite estimates to draw a data-driven conclusion that may use further as a reference in future studies. Many studies across the world have evaluated the reliability of Sentinel 5P pollution data with ground monitored measurements. Lorente et al. (2019) have examined the reliability of Sentinel TROPOMI tropospheric column NO_2_ density with ground monitored (ground monitored NO_2_ boundary layer height over the Eiffel Tower was used in this purpose) data and found a very good agreement (R^2^ = 0.88) between the two estimates. Griffin et al., (2019) study on validating TROPOMI data with aircraft and surface in situ NO_2_ observations over the Canadian oil sands found that the TROPOMI vertical NO_2_ column densities are strongly correlated (R^2^ = 0.86) with the aircraft and ground in situ NO_2_ observations with a low bias (15–30%).

### 4.2 Anthropogenic emission and ecosystem services

In this study, the spatial and temporal distribution and changes in different air pollution were measured for different cities across the world. A fixed timeframe (1^st^ February to 11^th^ May) was considered for the spatial and temporal analysis and subsequent interpretation. For all the 20 cities, NO_2_ concentration was found to be decreased with mixed intensities. Due to the imposition of worldwide lockdown and resulted in anthropogenic emission switch-off, air pollution across the world has been reduced significantly. Among the countries, the highest NO_2_ reduction was observed for Netherlands (70%), Japan (64%), Macao (60%), Lebanon (55%), Italy (54%), India (54%), Monaco (54%), North Korea (51%), Hungary (50%), and Kuwait (50%), respectively. While an incremental trend of NO_2_ emission was found in the Island countries, i.e., Kiribati (213%), Howland Island (136%), Jarvis Island (129%), Nauru (93%), Pacific Islands (Palau) (81%) along with other countries such as Indonesia (74%), Nepal (57%), Mozambique (56%), Norfolk Island (55%), and Jan Mayen (52%), where COVID1–9 lockdown has not implemented or followed strictly (**Fig. S11, Table. S2, Table. S3**). For CO, the maximum reduction was observed in Ecuador (6%), Colombia (6%), Venezuela (4%), Macau (4%), South Korea (4%), North Korea (4%), Byelarus (3%), Singapore (3%), Estonia (3%), and Latvia (3%), respectively. While, during the lockdown period, an increasing trend of CO emission was documented for some countries, such as Sao Tome and Principe (14%), Equatorial Guinea (14%), Gabon (13%), Argentina (13%), Falkland Islands (13%), Uruguay (12%), Congo (12%), Bouvet Island (11%), and Cameroon (11%), respectively. For both NO_2_ and CO, the maximum reduction is recorded for the countries which have been strongly affected by the COVID pandemic. The economic loss due to this exceeding level of air pollution has also been evaluated in this study. However, in this study, only the median externality values of the air pollutants are considered for the valuation and subsequent interpretation. This one dimension and linear valuation approach will not be able to track down the overall economic impact of air pollution on human life. Therefore, research that broadens the scope of valuation needs to be initiated for exploring the importance of proper monetary valuation in environmental studies.

### 4.3 Human mobility and its association with air pollution

The connection between human mobility and air pollution levels in selected cities were also examined in this research. Both Apple and Google mobility data were used for this purpose. Results derived from both the report suggest that due to the mandatory lockdown and resulted in limited outdoor human activities, mobility has been reduced significantly across the world. This drastic reduction of human mobility could contribute to the reduced level of air pollution observed in the last few months. For most of the cities considered in this study, human mobility has been reduced up to 80% from the baseline mobility. The highest reduction in mobility was found in the European cities. To prevent infection, the authorities in these cities implemented preventive measures, which included partial lockdown in different sectors, including restricted outdoor social activities. This mandatory imposition of lockdown has resulted in a reduced level of traffic volume in cities (**Fig. 11, Table. S6**). The mobility analysis thus suggests that by introducing sustainable transport plans and policies, air pollution in the urban regions can be minimised to a certain extent. The periodic and temporary lockdown can also be adopted in the highly polluted cities if no other alternatives are feasible at the place. A similar strategy has already been adopted by New Delhi Government by introducing “odd/even” transport scheme where private vehicles with odd digit (1, 3, 5, 7, 9) registration numbers will be allowed on roads on odd dates and vehicles with even digit (0, 2, 4, 6, 8) registration numbers can use the vehicles on even dates. In addition, the Mahato et al. study has observed a 40% to 50% improvement in air quality in Delhi within the first week of lockdown. He et al. (2020) study on short-term impacts of COVID-19 lockdown on urban air pollution has found that within a week, the AQI in the locked-down cities in China has been reduced by 19.84 points (PM_2.5_ goes down by 14.07 µg m^−3^) compared to the cities where lockdown has not been implemented strictly. The findings suggest an increased clean air ecosystem services in cities under the cessation of human activities.

## 5. Conclusion

This study has evaluated the effect of COVID-19 lockdown on air quality ecosystem services across the world. A total of 20 major cities were considered for the analysis and subsequent interpretation. Both satellite and ground air pollution data were utilised for examining the association between COVID pandemic led lockdown and improving status of air quality ecosystem services across the cities. The major findings of this research are:

1. Among the 20 cities, the average NO_2_ concentration (1 Feb to 11 May) was found highest in Tehran, followed by Milan, New York, Paris, Turin, Chicago, Cologne, and Philadelphia.
2. The lowest NO_2_ concentration (1 Feb to 11 May) was observed in Sao Paulo, Brussels, and Denver.
3. For NO_2_, the highest reduction was detected in Paris (45.94%), followed by Detroit (40.29%), Milan (36.85%), Turin (36.83%), Frankfurt (36.36%), Philadelphia(34.45%), London (34.15%), and Madrid (34.03%), respectively.
4. While, a comparably lower reduction of NO_2_ is observed in Los Angeles (10.54%), Sao Paulo (17.17%), Antwerp (24.14%), Tehran (24.54%), and Rotterdam (26.72%), during the lockdown period.
5. For CO, the maximum reduction was recorded for New York (4.24%), followed by Detroit (4.09%), Sao Paulo (3.88%), Philadelphia (3.45%), Milan (3.17%), Barcelona(2.86%), respectively.
6. The daily NO_2_ and SO_2_ AQI during the lockdown period suggest that all the cities are benefitted by having good quality air due to anthropogenic pollution switch-off and restricted human interventions.
7. Among the cities, the highest economic values (derived from public health burden valuation approach) was estimates for New York (501 million US$), followed by London (375 million US$), Chicago (137 million US$), Paris (124 million US$), Madrid (90 million US$), Philadelphia (89 million US$), Milan (78 million US$), Cologne (67 million US$), Los Angeles (67 million US$), Frankfurt (52 million US$), Turin (45 million US$), Detroit (43 million US$), Barcelona (41 million US$), Sao Paulo (40 million US$), Tehran (37 million US$), Denver (30 million US$), Antwerp (16 million US$), Utrecht (14 million US$), Brussels (9 million US$), and Rotterdam (9 million US$), respectively.
8. For NO_2_, the economic significance of reduced anthropogenic emission is found maximum in Tehran (31700 $), followed by London (21887 $), New York (12975 $), and Madrid (9072 $).
9. For CO, the maximum ecosystem service value was calculated maximum for Sao Paulo (42302 $), followed by New York (36478 $), London (17043 $), Detroit (16038 $), and Los Angeles (14472 $).
10. Among the countries, the highest NO_2_ reduction was observed for Netherlands (70%), Japan (64%), Macao (60%), Lebanon (55%), Italy (54%), India (54%), Monaco (54%), North Korea (51%), Hungary (50%), and Kuwait (50%).
11. For CO, the maximum reduction was observed in Ecuador (6%), Colombia (6%), Venezuela (4%), Macau (4%), South Korea (4%), North Korea (4%), Byelarus (3%), Singapore (3%), Estonia (3%), and Latvia (3%).

The present research has made an effort to investigate the human impact on the natural environment by taking COVID-19 lockdown and its resultant reduction of air pollution. Both physical and monetary valuation was carried out to assess the synergic effect of this pandemic led lockdown on air pollutions at 20 cities across the world. A strong connection between human interventions and accelerating levels of air pollution was observed in most of these cities. Both satellite and ground-based estimates are suggesting the positive effect of the limited human interference on natural environments. Further research in this direction is needed to explore this synergic association more explicitly.

## Data Availability

All data such European Space Agency (ESA) Sentinel satellite data, EPA pollution data, and Google and Apple mobility data, etc. used this article are open and free accessible.

---

**Fig. S1.**
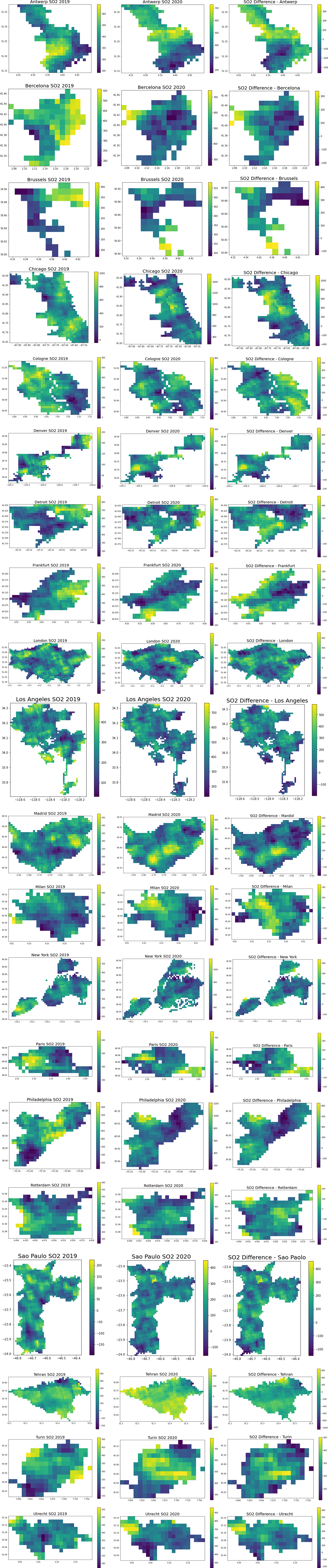
Spatial distribution of SO_2_ (μmol m^−2^) in the selected cities in 2019 and 2020 (from Feb to May). Spatial maps in third panel shows the spatial difference in SO_2_ concentration between 2019 and 2020.

**Fig. S2.**
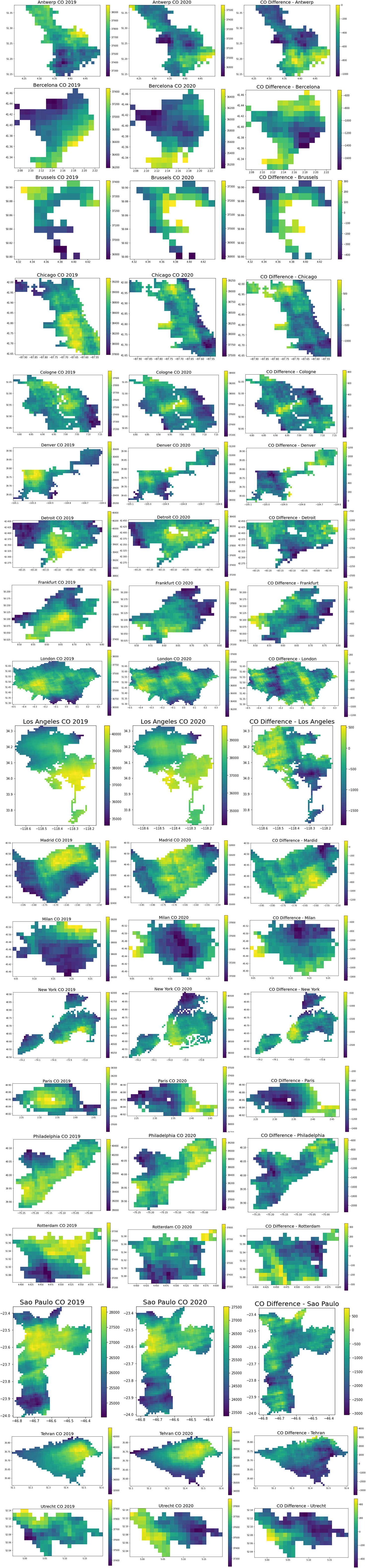
Spatial distribution of CO (μmol m^−2^) in the selected cities in 2019 and 2020 (from Feb to May). Spatial maps in third panel shows the spatial difference in CO concentration between 2019 and 2020.

**Fig. S3.**
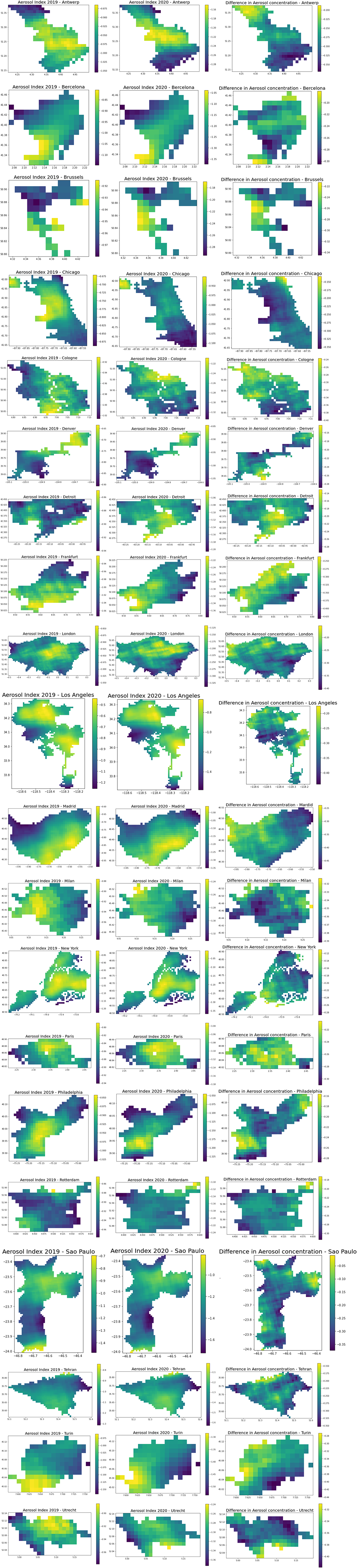
Spatial distribution of Aerosol index in the selected cities in 2019 and 2020 (from Feb to May). Spatial maps in third panel shows the spatial difference in aerosol concentration between 2019 and 2020.

**Fig. S4.**
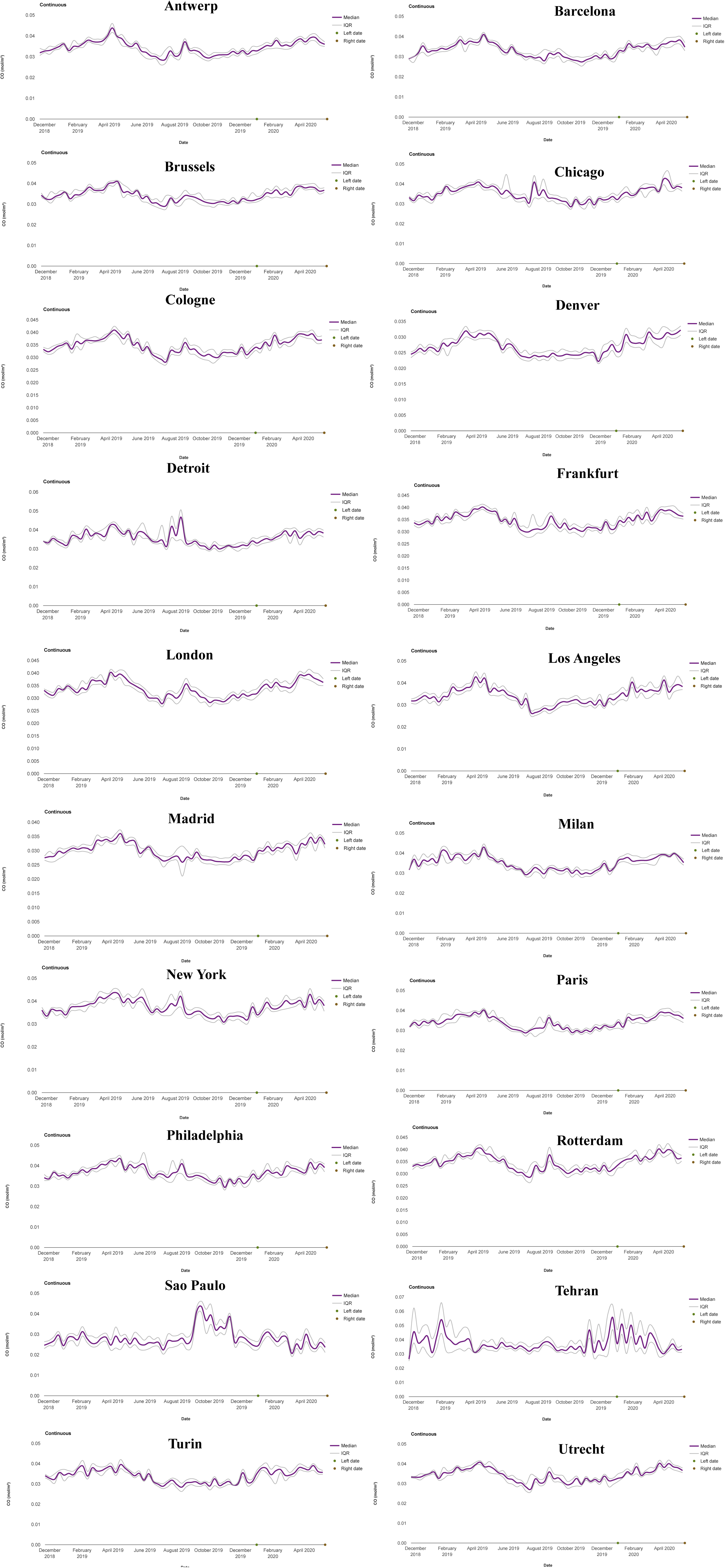
Temporal variation of CO (μmol m^−2^) concentration in the selected cities from August 2018 to May 2020 derived from Sentinel 5P TROPOMI data.

**Fig. S5.**
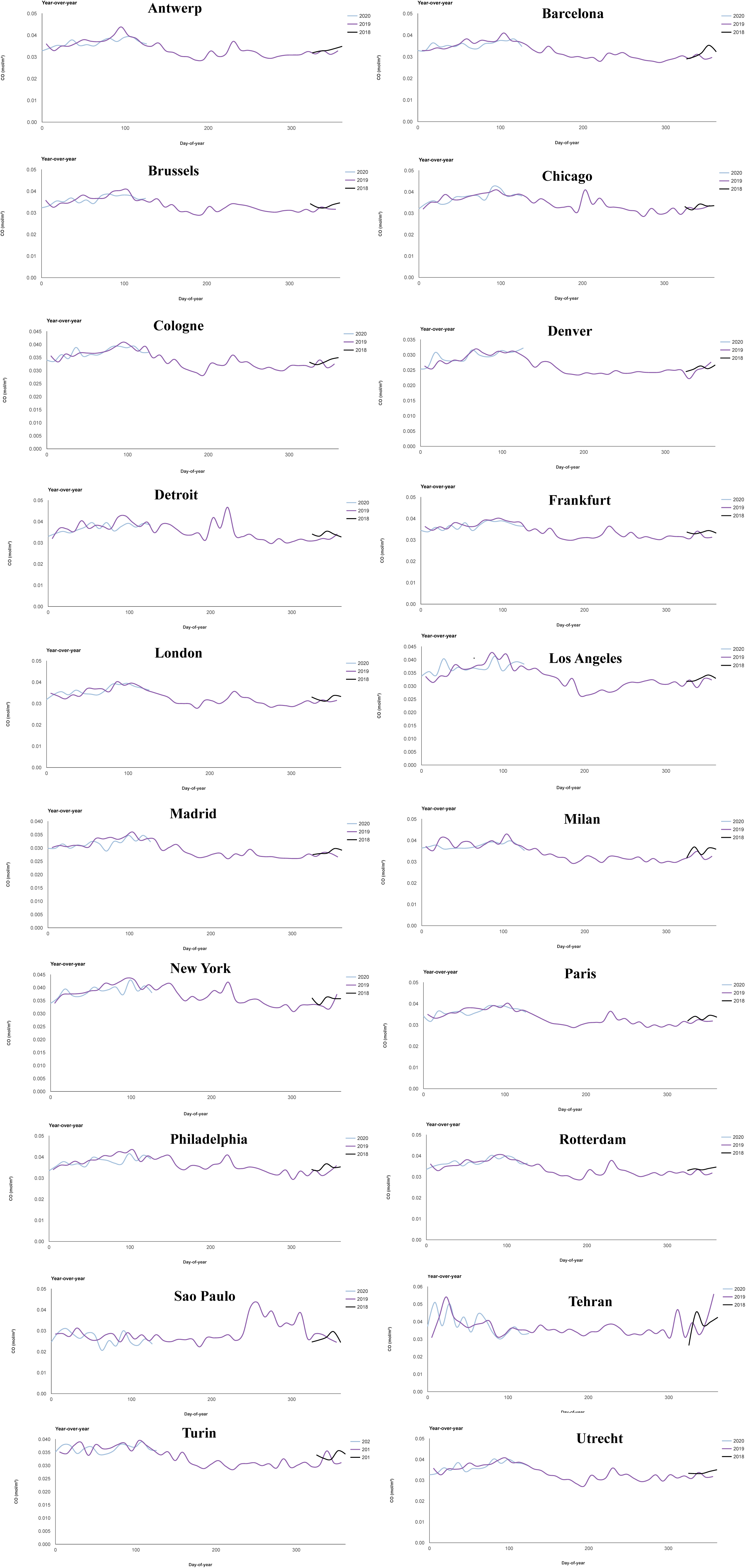
Temporal variation of CO (μmol m^−2^) concentration in the selected cities in 2018, 2019, and 2020 derived from Sentinel 5P TROPOMI data.

**Fig. S6.**
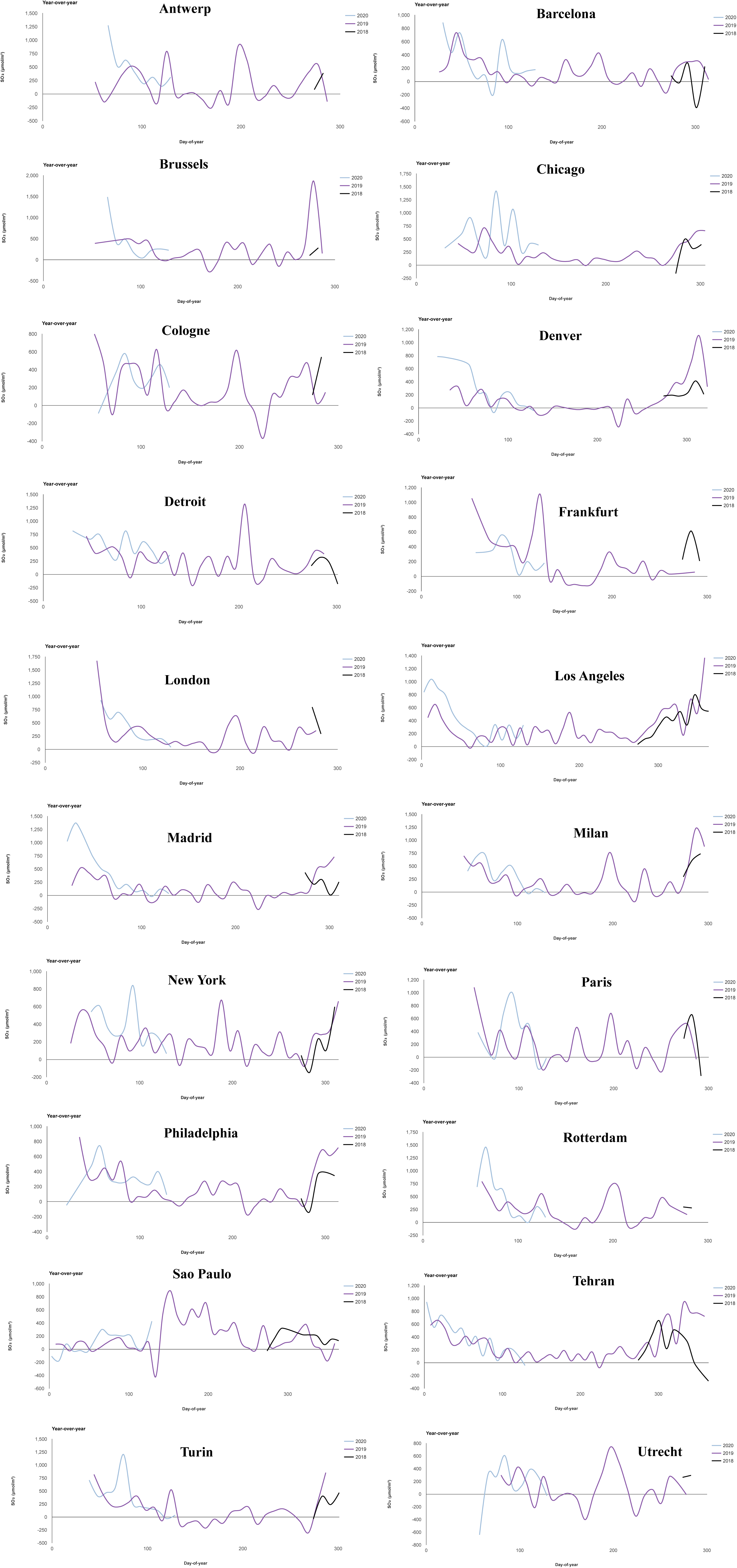
Temporal variation of SO_2_ (μmol m^−2^) concentration in the selected cities from August 2018 to May 2020 derived from Sentinel 5P TROPOMI data.

**Fig. S7.**
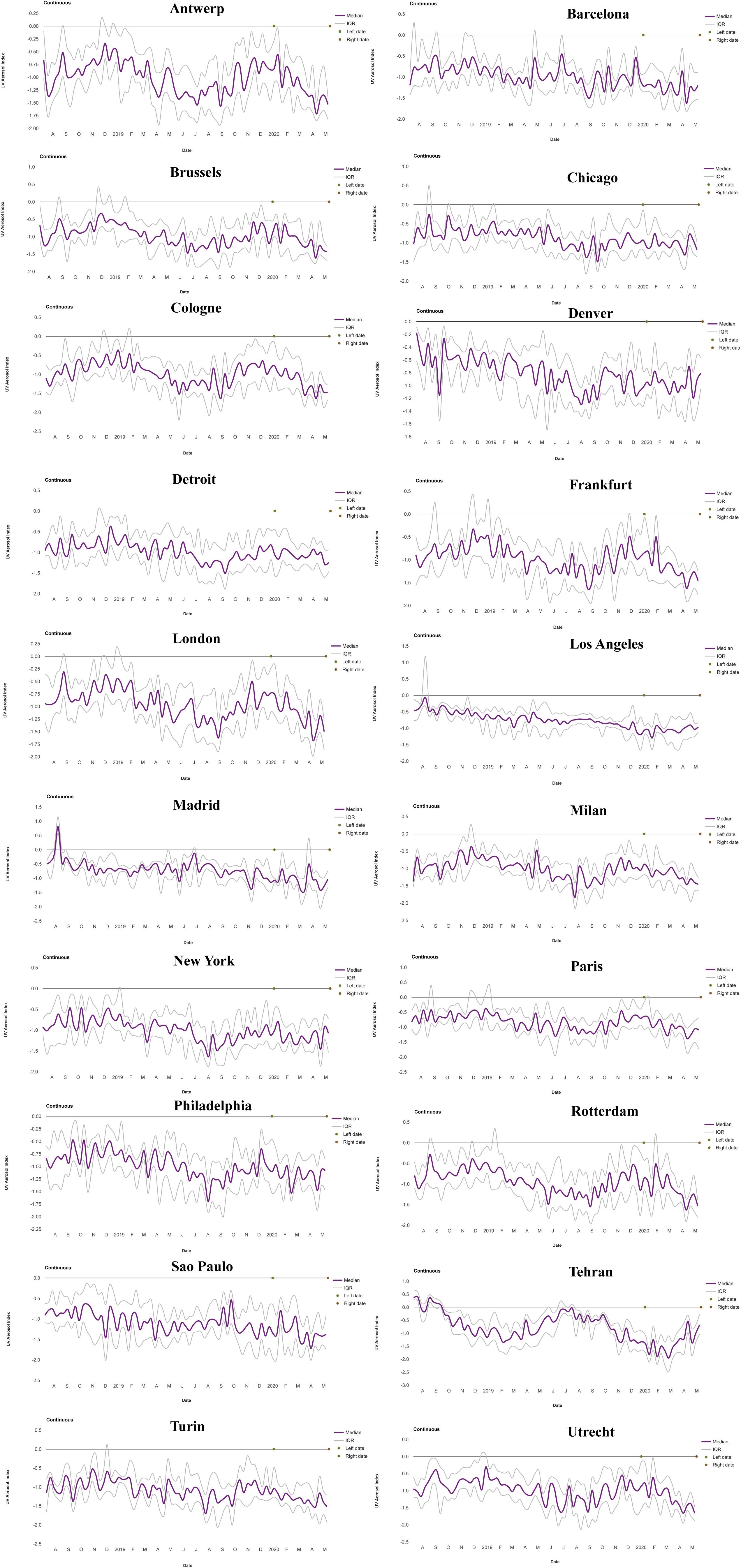
Temporal variation of aerosol concentration in the selected cities from August 2018 to May 2020 derived from Sentinel 5P TROPOMI observation.

**Fig. S8.**
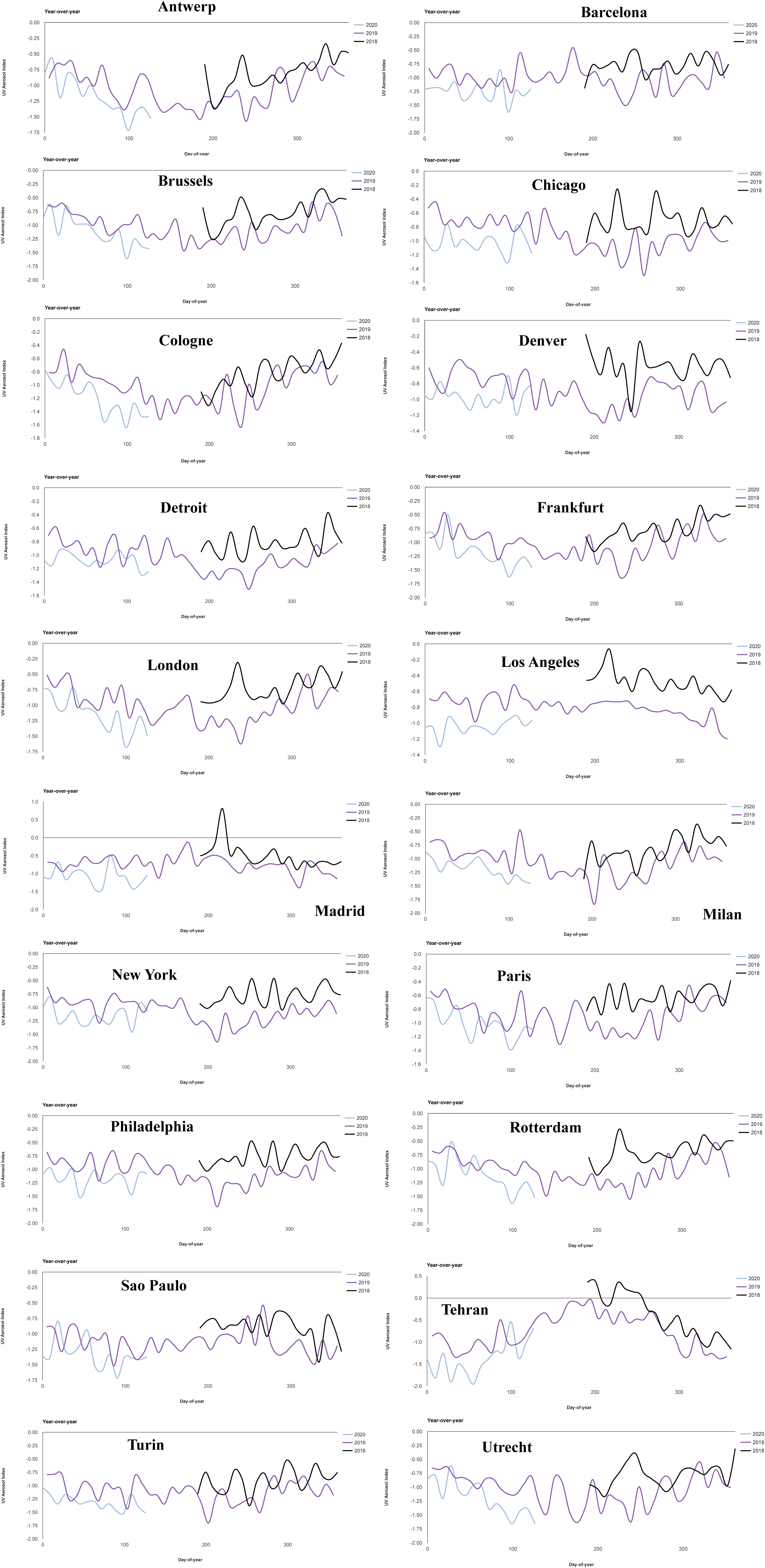
Temporal variation of aerosol concentration in the selected cities in 2018, 2019, and 2020 derived from Sentinel 5P TROPOMI observation.

**Fig. S9.**
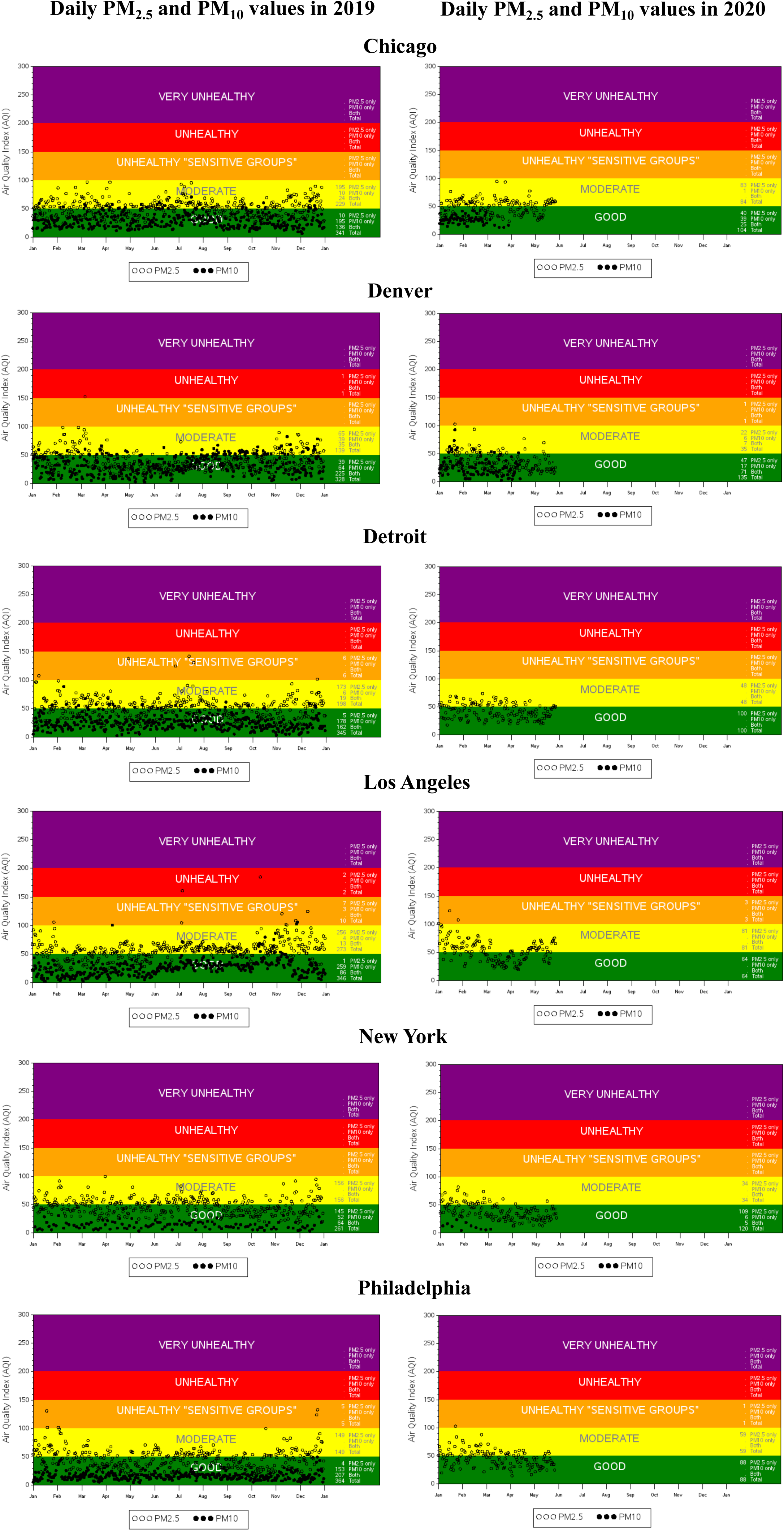
Shows the ground monitored air quality index (based on PM_2.5_ and PM_10_) in 2019 and 2020 in the selected cities.

**Fig. S10.**
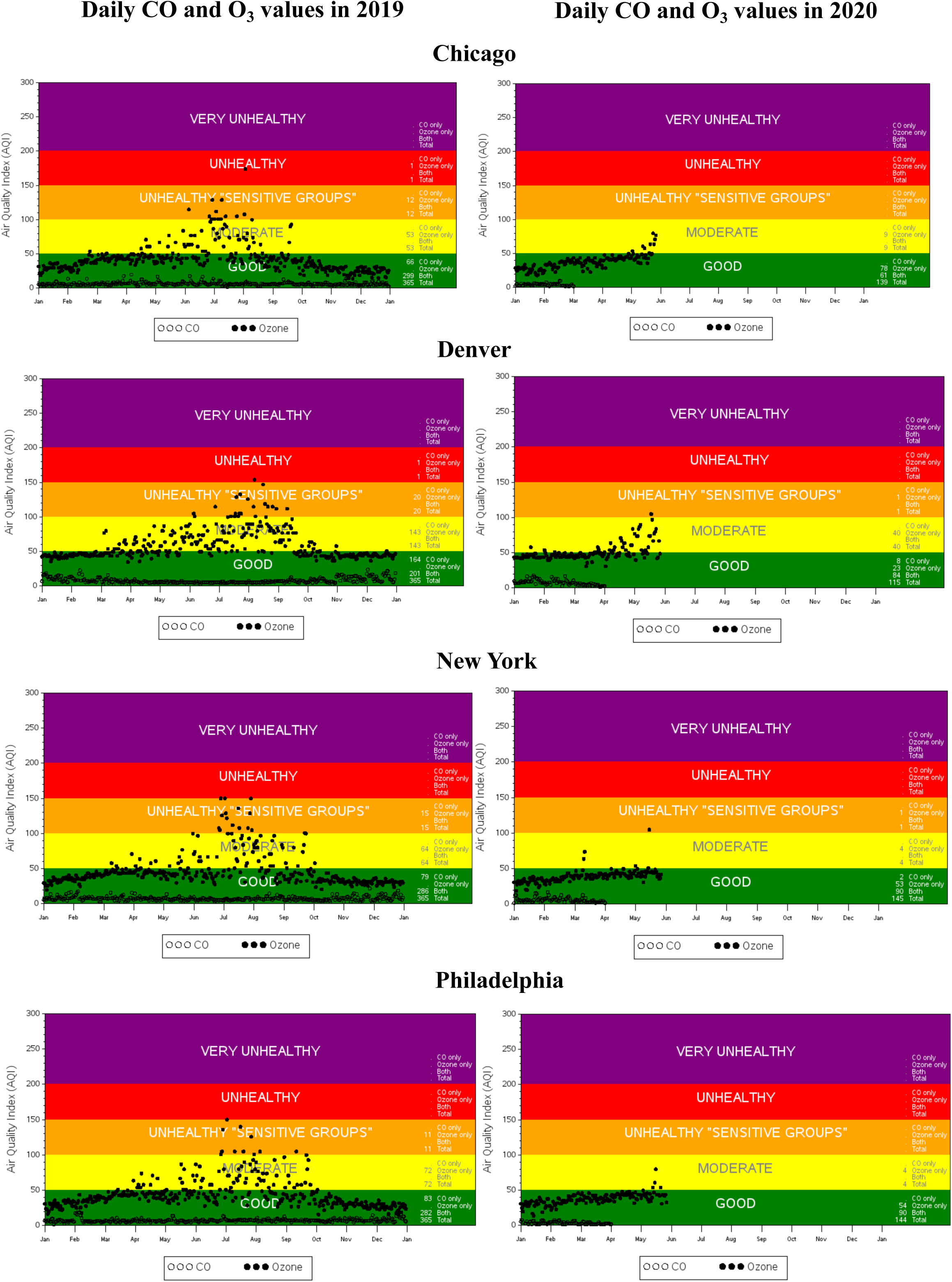
Shows the ground monitored air quality index (based on CO and O3) in 2019 and 2020 in the selected cities.

**Fig. S11.**
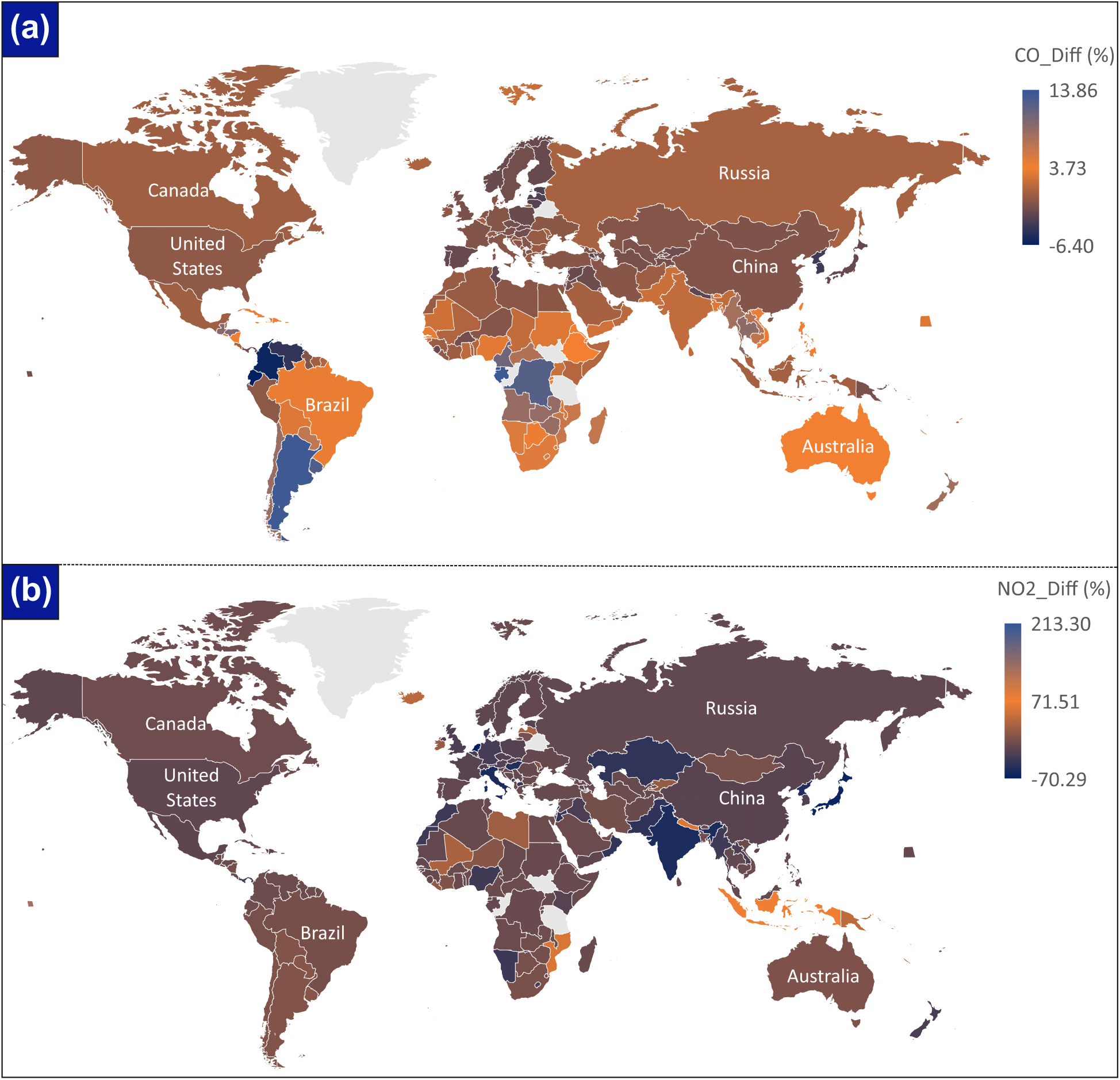
Changes in NO_2_ and CO concentration during the study period (1st Feb to 11th May in 2019 and 2020). NO2 changes are maximum in few Asian countries and European countries. Whereas, CO chnages are promnent in China, USA, and few European countries.

**Fig. S12.**
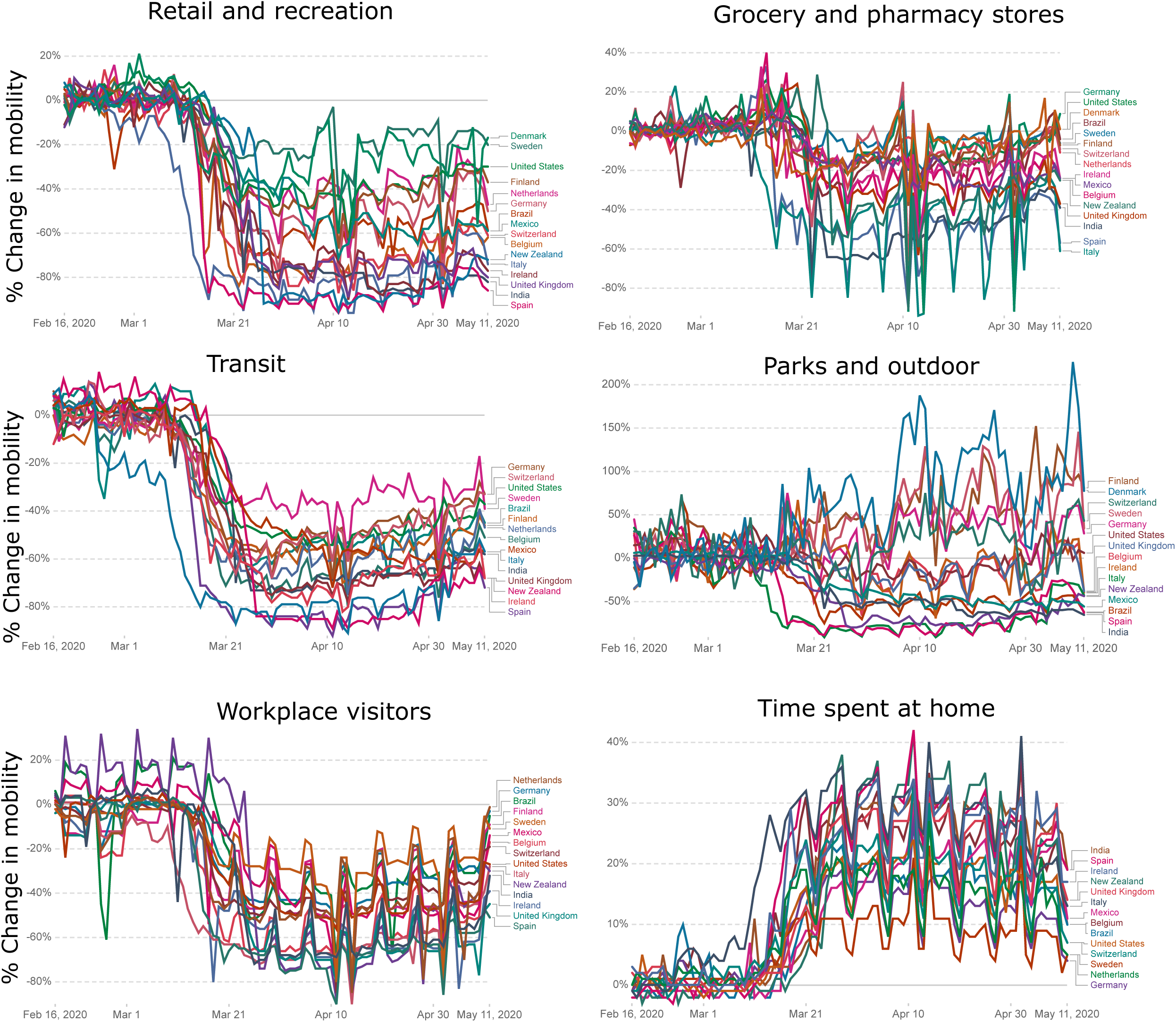
Changes in human mobility observed during the lock down period. Six mobility factors, i.e.., retial and recreation, grocery and pharmacy, transit, parks and outdoor, workplace visitors, and time spent at home is evaluated in this study.

**Fig. S13.**
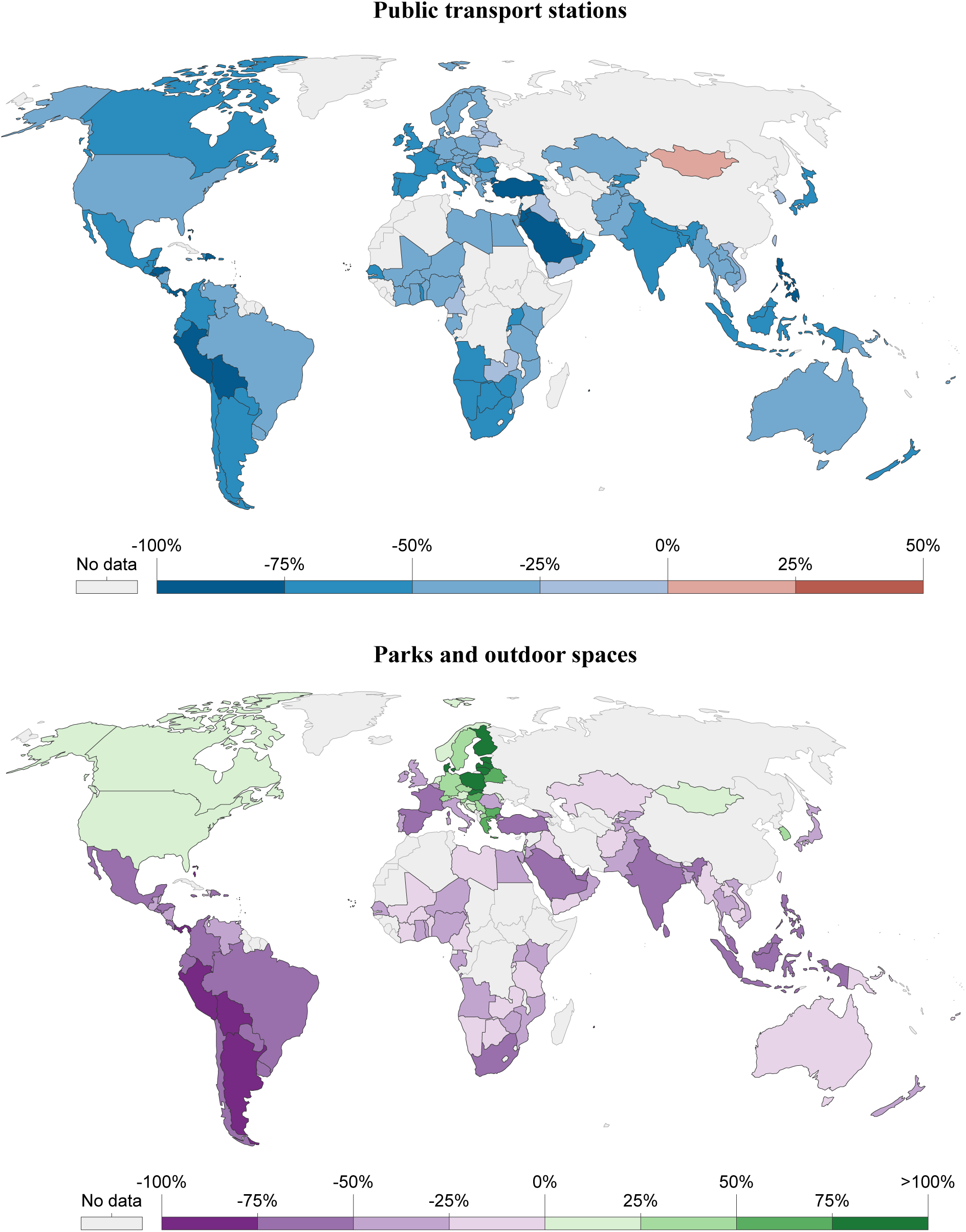
Spatial variability of public transport and parks/outdoor mobility during the lockdown period.

**Fig. S14.**
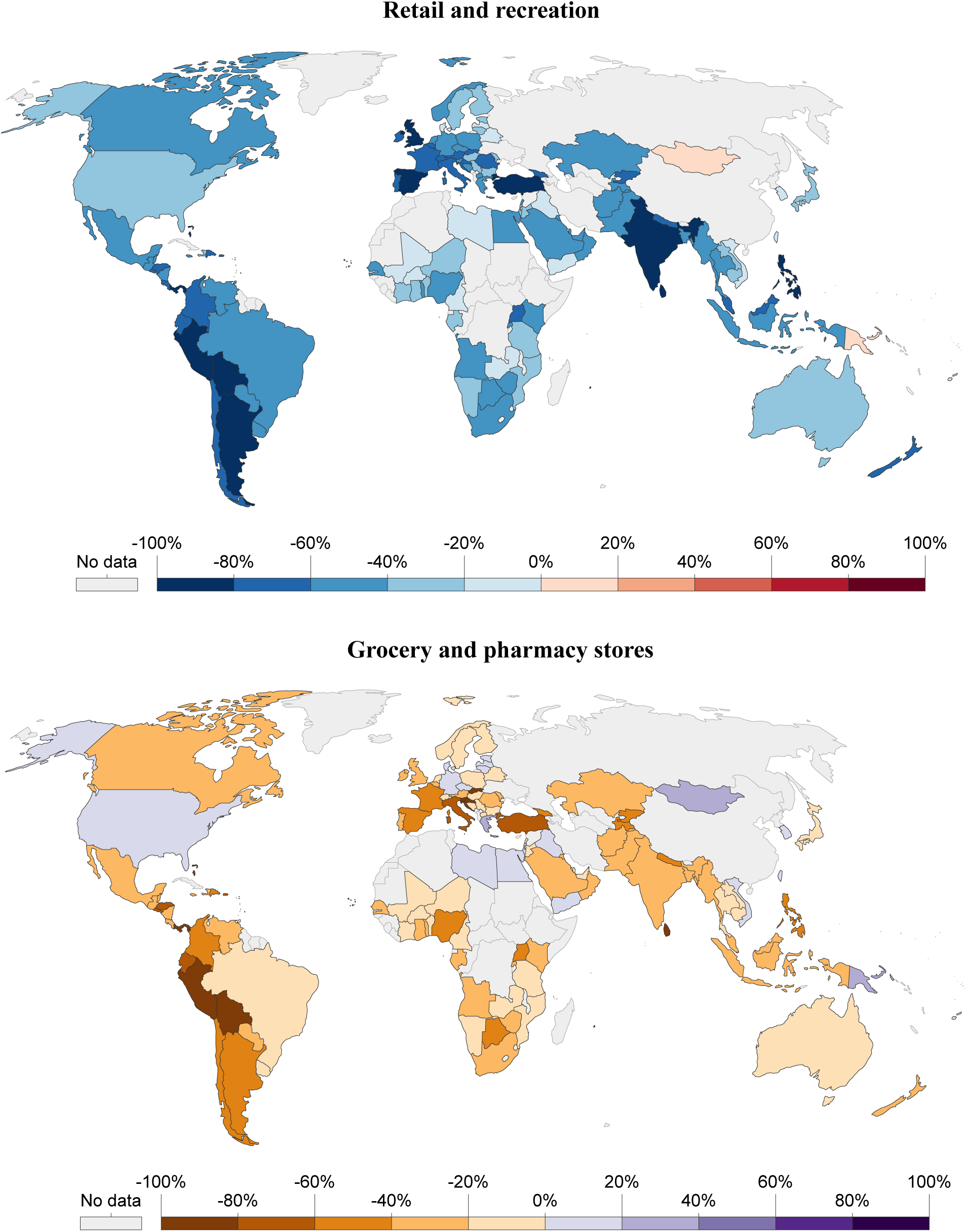
Spatial variability of retail/recreation and grocery/pharmacy mobility during the lock-down period.

**Fig. S15.**
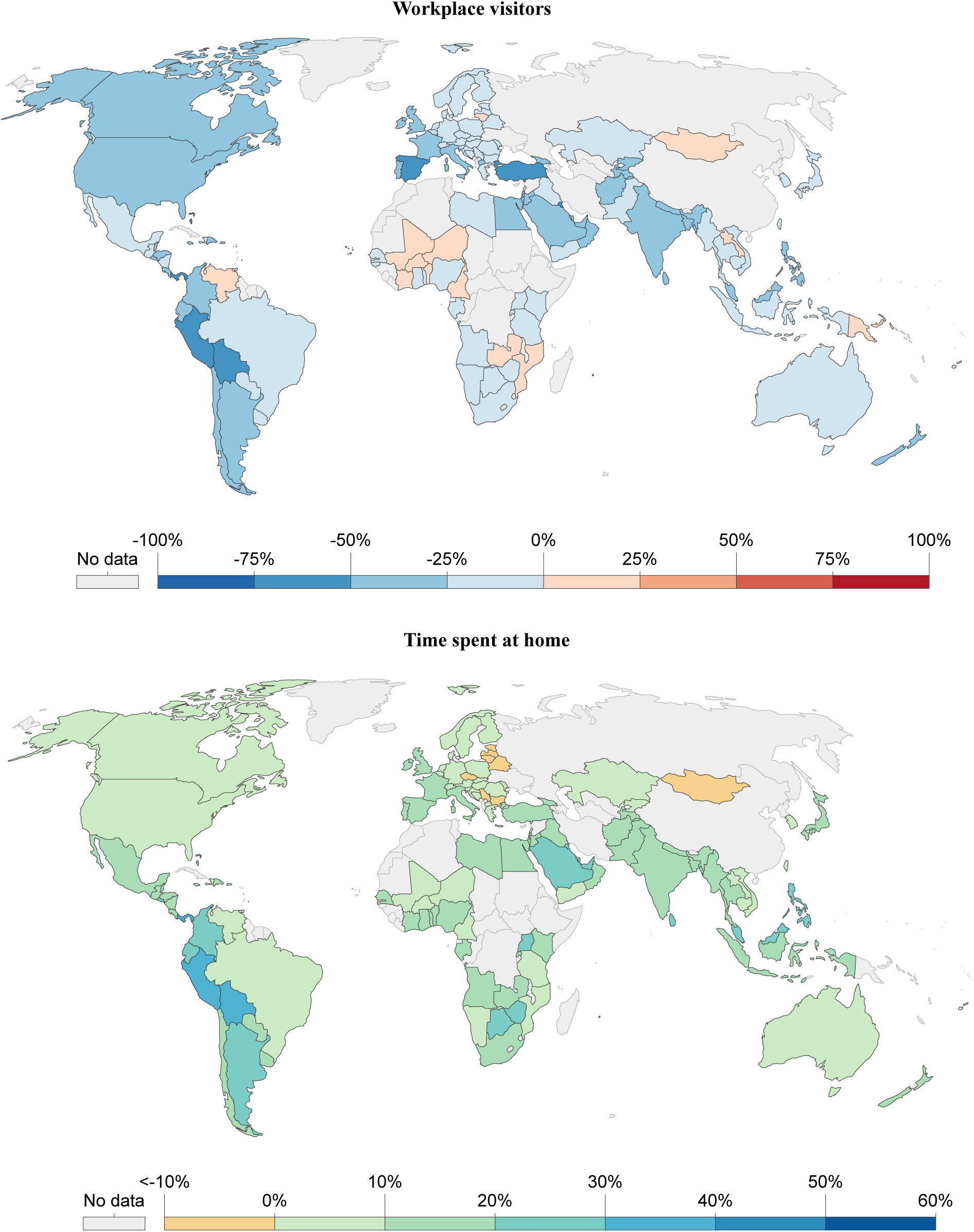
Spatial variability of workplace and residential mobility during the lockdown period.

**Table. S1.** AQI categorization for different air pollutants.

| AQI | NO<sub>2</sub><br>(ppb) | CO<br>(ppm) | O<sub>3</sub><br>(ppm/hr) | PM<sub>10</sub><br>(µg m<sup>-3</sup>) | PM<sub>2.5</sub><br>(µg m<sup>-3</sup>) | SO<sub>2</sub><br>(ppb) |
| --- | --- | --- | --- | --- | --- | --- |
| Good | <=53 | <=4.4 | <=0.054 | <=54 | <=12 | <=35 |
| Moderate | 54-100 | 4.5-9.4 | 0.055-0.070 | 55-154 | 12.1-35.4 | 36-75 |
| Unhealthy | 101-360 | 9.5-12.4 | 0.071-0.085 | 155-254 | 35.5-55.4 | 76-185 |
| Unhealthy | 361-649 | 12.5-15.4 | 0.086-0.105 | 255-354 | 55.5-150.4 | 186-304 |
| Very | 650-1,249 | 15.5-30.4 | 0.106-0.200 | 355-424 | 150.5-250.4 | 305-604 |
| Hazardous | >=1,250 | >=30.5 | >=0.405 | >=425 | >=250.5 | >=605 |

**Table. S2.** Change statistics of NO2 during the study period (Feb 1 to May 11). Minus and plus signs are indicating reduction and increases of NO2.

| Country | ΔNO<sub>2</sub> | Country | $\Delta$ NO<sub>2</sub> |
| --- | --- | --- | --- |
| Netherlands | -70.29 | Kiribati | 213.30 |
| Japan | -63.99 | Howland Island | 135.82 |
| Macau | -59.68 | Jarvis Island | 128.79 |
| Man, Isle of | -57.54 | Nauru | 93.02 |
| Lebanon | -54.75 | Pacific Islands (Palau) | 80.82 |
| Italy | -54.41 | Indonesia | 74.39 |
| India | -53.68 | Nepal | 56.72 |
| Monaco | -53.63 | Mozambique | 56.19 |
| North Korea | -50.84 | Norfolk Island | 54.61 |
| Hungary | -50.49 | Jan Mayen | 52.15 |
| Kuwait | -49.97 | Mayotte | 48.85 |
| Pakistan | -42.58 | New Caledonia | 41.76 |
| Kazakhstan | -41.84 | Papua New Guinea | 40.68 |
| Oman | -41.42 | Iceland | 37.63 |
| Jordan | -40.51 | Juan De Nova Island | 37.59 |
| Macedonia | -37.29 | Niue | 29.18 |
| Namibia | -35.05 | Mali | 28.36 |
| Liechtenstein | -33.81 | Latvia | 28.05 |
| Morocco | -33.57 | Midway Islands | 25.03 |
| Myanmar (Burma) | -32.59 | Maldives | 22.78 |
| Nigeria | -32.39 | Libya | 21.53 |
| Montenegro | -31.59 | Ireland | 17.43 |
| Singapore | -29.89 | Kyrgyzstan | 16.11 |
| Germany | -29.82 | Montserrat | 15.18 |
| Denmark | -29.35 | Marshall Islands | 14.28 |
| Panama | -27.38 | Liberia | 13.72 |
| Laos | -27.14 | Paraguay | 9.26 |
| Iraq | -26.96 | Uruguay | 8.80 |
| New Zealand | -26.94 | Niger | 8.67 |
| Jersey | -26.14 | Pitcairn Islands | 8.42 |

**Table. S3.** Change statistics of CO during the study period (Feb 1 to May 11). Minus and plus signs are indicating reduction and increases of CO.

| Country | ΔCO | Country | ΔCO |
| --- | --- | --- | --- |
| Ecuador | -6.40 | Sao Tome and Principe | 13.86 |
| Colombia | -5.90 | Equatorial Guinea | 13.68 |
| Venezuela | -4.32 | South Georgia | 13.53 |
| Macau | -4.09 | Gabon | 13.27 |
| South Korea | -3.71 | Argentina | 13.05 |
| North Korea | -3.70 | Falkland Islands (Islas Malvinas) | 12.64 |
| Byelarus | -3.27 | Uruguay | 12.15 |
| Singapore | -3.10 | Congo | 11.88 |
| Estonia | -3.06 | Bouvet Island | 11.36 |
| Latvia | -2.93 | Heard Island & McDonald Islands | 11.25 |
| Malta | -2.84 | Cameroon | 10.56 |
| Lithuania | -2.77 | Honduras | 9.70 |
| Aruba | -2.74 | French Southern & Antarctic Lands | 9.53 |
| Man, Isle of | -2.57 | Guatemala | 9.30 |
| Nepal | -2.53 | Zaire | 9.22 |
| Armenia | -2.46 | Thailand | 9.04 |
| Portugal | -2.30 | Zambia | 8.66 |
| Tunisia | -2.22 | Angola | 8.57 |
| Jersey | -2.21 | Zimbabwe | 8.53 |
| Andorra | -2.20 | Chile | 8.39 |
| Japan | -2.15 | Glorioso Islands | 8.39 |
| St. Pierre and Miquelon | -2.07 | Norfolk Island | 8.08 |
| Finland | -2.06 | New Zealand | 7.94 |
| Syria | -2.05 | Myanmar (Burma) | 7.81 |
| Spain | -2.03 | Belize | 7.80 |
| Sierra Leone | -1.99 | Reunion | 7.64 |
| Norway | -1.98 | Mauritius | 7.35 |
| Poland | -1.98 | Central African Republic | 7.22 |
| Jan Mayen | -1.91 | Guadeloupe | 6.98 |
| Iraq | -1.85 | Laos | 6.93 |

**Table. S4.**
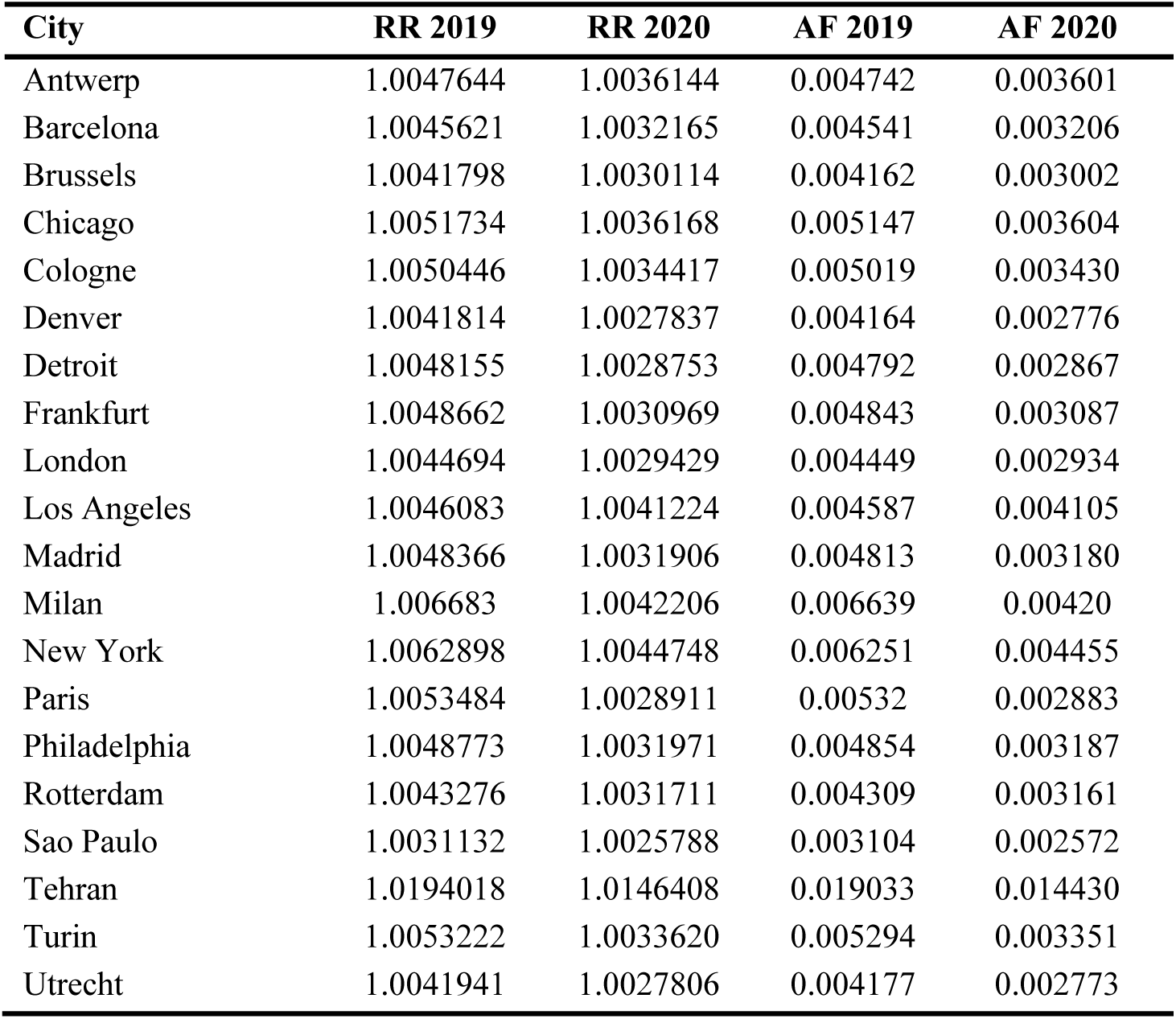
Summary statistics of relative risk (RR) and attributable fraction (AF) in 2019 and 2020.

**Table. S5.** Summary statistics of health burden and economic burden of 20 major cities. CV HB = Cardiovascular health burden, CRD GB = Chronic respiratory disease health burden, THB = total health burden, VSL = value of statistical life (million US$), EB = economic burden (million US$).

| City | CV HB<br>2019 | CRD HB<br>2019 | THB<br>2019 | CV HB<br>2020 | CRD HB<br>2020 | THB<br>2020 | VSL | EB<br>2019 | EB<br>2020 |
| --- | --- | --- | --- | --- | --- | --- | --- | --- | --- |
| Antwerp | 7 | 1 | 8 | 5 | 1 | 6 | 8 | 67 | 51 |
| Barcelona | 21 | 6 | 27 | 15 | 4 | 19 | 5 | 138 | 97 |
| Brussels | 3 | 1 | 4 | 2 | 0 | 3 | 8 | 31 | 22 |
| Chicago | 37 | 8 | 45 | 26 | 6 | 31 | 10 | 456 | 320 |
| Cologne | 22 | 3 | 25 | 15 | 2 | 17 | 8 | 213 | 145 |
| Denver | 7 | 2 | 9 | 5 | 1 | 6 | 10 | 89 | 59 |
| Detroit | 9 | 2 | 10 | 5 | 1 | 6 | 10 | 106 | 64 |
| Frankfurt | 15 | 2 | 17 | 10 | 1 | 11 | 8 | 144 | 92 |
| London | 110 | 29 | 139 | 72 | 19 | 92 | 8 | 1102 | 727 |
| Los Angeles | 51 | 11 | 62 | 46 | 10 | 56 | 10 | 634 | 568 |
| Madrid | 40 | 11 | 51 | 27 | 7 | 34 | 5 | 267 | 176 |
| Milan | 31 | 4 | 35 | 20 | 3 | 22 | 6 | 211 | 134 |
| New York | 140 | 31 | 171 | 100 | 22 | 122 | 10 | 1744 | 1243 |
| Paris | 32 | 4 | 36 | 17 | 2 | 20 | 7 | 270 | 146 |
| Philadelphia | 21 | 5 | 25 | 14 | 3 | 17 | 10 | 258 | 169 |
| Rotterdam | 3 | 1 | 4 | 2 | 1 | 3 | 9 | 35 | 26 |
| Sao Paulo | 103 | 28 | 130 | 85 | 23 | 108 | 2 | 234 | 194 |
| Tehran | 117 | 11 | 127 | 88 | 8 | 97 | 1 | 152 | 115 |
| Turin | 18 | 2 | 20 | 11 | 2 | 13 | 6 | 123 | 78 |
| Utrecht | 4 | 1 | 5 | 2 | 1 | 3 | 9 | 41 | 27 |

**Table. S6.** Changes in human mobility (%) from the baseline (mobility on 13^th^ January) during the lockdown period (1^st^ February to 11^th^ May 2020).

| City | Jan (From 13<sup>th</sup>) |  | Feb |  | Mar |  | April |  | May (up to 11) |  |
| --- | --- | --- | --- | --- | --- | --- | --- | --- | --- | --- |
|  | Driving | Transit | Driving | Transit | Driving | Transit | Driving | Transit | Driving | Transit |
| Antwerp | 14.00 | 4.39 | 20.80 | 23.94 | -31.05 | -35.98 | -58.79 | -75.84 | -47.34 | -66.90 |
| Barcelona | 8.60 | 4.47 | 18.15 | 63.81 | -44.91 | 4.86 | -85.04 | -88.10 | -74.71 | -79.85 |
| Brussels | 9.49 | 14.53 | 15.07 | 32.10 | -37.64 | -39.03 | -65.32 | -81.19 | -52.91 | -73.32 |
| Chicago | 5.61 | -0.53 | 12.73 | 4.47 | -18.38 | -39.60 | -41.98 | -77.76 | -23.97 | -74.56 |
| Cologne | -4.19 | -4.63 | -1.08 | 43.30 | -37.46 | -17.98 | -51.98 | -55.32 | -35.83 | -50.94 |
| Denver | 5.34 | -1.19 | 6.72 | 0.61 | -24.10 | -36.10 | -48.45 | -70.08 | -28.47 | -64.71 |
| Frankfurt | 4.28 | ----- | 5.72 | ----- | -30.92 | ----- | -44.89 | ----- | -32.83 | ----- |
| London | 10.85 | 11.89 | 14.61 | 17.76 | -26.71 | -38.02 | -67.16 | -86.27 | -60.17 | -82.80 |
| Los Angeles | 12.41 | 3.30 | 17.30 | 7.81 | -22.80 | -39.09 | -51.15 | -76.52 | -34.31 | -72.70 |
| Madrid | 9.60 | 9.69 | 16.22 | 14.44 | -52.45 | -58.34 | -84.25 | -93.47 | -72.18 | -88.56 |
| Milan | ----- | 9.96 | ----- | 6.19 | ----- | -70.30 | ----- | -82.30 | ----- | -65.81 |
| New York | 4.17 | -2.28 | 8.47 | 1.30 | -26.78 | -48.74 | -54.87 | -86.43 | -38.78 | -83.47 |
| Paris | -8.05 | 0.83 | -15.30 | 11.26 | -57.52 | -49.32 | -82.96 | -89.61 | -75.31 | -83.91 |
| Philadelphia | 4.19 | -6.64 | 9.98 | -4.16 | -20.88 | -38.65 | -43.17 | -71.29 | -23.61 | -69.32 |
| Rotterdam | 6.34 | 4.90 | 4.70 | 8.20 | -30.60 | -40.12 | -44.29 | -67.66 | -33.26 | -61.54 |
| Sao Paulo | 4.51 | -0.97 | 12.51 | 4.88 | -28.66 | -35.99 | -61.68 | -81.04 | -57.29 | -80.84 |
| Utrecht | 0.44 | -1.09 | -0.58 | 6.40 | -35.94 | -45.48 | -51.19 | -72.12 | -41.26 | -66.09 |

## References

Abhijith, K. V, Kumar, P., Gallagher, J., McNabola, A., Baldauf, R., Pilla, F., Broderick, B., Di Sabatino, S., Pulvirenti, B., 2017. Air pollution abatement performances of green infrastructure in open road and built-up street canyon environments – A review. Atmos. Environ. 162, 71–86. https://doi.org/10.1016/j.atmosenv.2017.05.014

Adekola, O., Mitchell, G., Grainger, A., 2015. Inequality and ecosystem services: The value and social distribution of Niger Delta wetland services. Ecosyst. Serv. 12, 42–54. https://doi.org/10.1016/j.ecoser.2015.01.005

Affek, A.N., Kowalska, A., 2017. Ecosystem potentials to provide services in the view of direct users. Ecosyst. Serv. 26, 183–196. https://doi.org/10.1016/j.ecoser.2017.06.017

Ash, N., Blanco, H., Garcia, K., & Brown, C. (2010). Ecosystems and human well-being: a manual for assessment practitioners. Island Press.

Bao, R., Zhang, A., 2020. Does lockdown reduce air pollution? Evidence from 44 cities in northern China. Sci. Total Environ. 731, 139052. https://doi.org/10.1016/j.scitotenv.2020.139052

Baró, F., Chaparro, L., Gómez-Baggethun, E., Langemeyer, J., Nowak, D.J., Terradas, J., 2014. Contribution of ecosystem services to air quality and climate change mitigation policies: The case of urban forests in Barcelona, Spain. Ambio 43, 466–479. https://doi.org/10.1007/s13280-014-0507-x

Bastian, O., Syrbe, R.-U., Rosenberg, M., Rahe, D., Grunewald, K., 2013. The five pillar EPPS framework for quantifying, mapping and managing ecosystem services. Ecosyst. Serv. 4, 15–24. https://doi.org/10.1016/j.ecoser.2013.04.003

Bherwani, H., Nair, M., Musugu, K., Gautam, S., Gupta, A., Kapley, A., Kumar, R., 2020. Valuation of air pollution externalities: comparative assessment of economic damage and emission reduction under COVID-19 lockdown. Air Qual. Atmos. Heal. 13, 683–694. https://doi.org/10.1007/s11869-020-00845-3

Borsdorff, T., Aan de Brugh, J., Hu, H., Aben, I., Hasekamp, O., Landgraf, J., 2018. Measuring Carbon Monoxide With TROPOMI: First Results and a Comparison With ECMWF-IFS Analysis Data. Geophys. Res. Lett. 45, 2826–2832. https://doi.org/10.1002/2018GL077045

Braat, L.C., de Groot, R., 2012. The ecosystem services agenda:bridging the worlds of natural science and economics, conservation and development, and public and private policy. Ecosyst. Serv. 1, 4–15. https://doi.org/10.1016/j.ecoser.2012.07.011

Burkhard, B., Kandziora, M., Hou, Y., Müller, F., 2014. Ecosystem service potentials, flows and demands-concepts for spatial localisation, indication and quantification. Landsc. Online 34, 1–32. https://doi.org/10.3097/LO.201434

Burkhard, B., Kroll, F., Müller, F., Windhorst, W., 2009. Landscapes’ capacities to provide ecosystem services – A concept for land-cover based assessments. Landsc. Online 15, 1–22. https://doi.org/10.3097/LO.200915

Castro, A., Künzli, N., Götschi, T., 2017. Health benefits of a reduction of PM10 and NO2 exposure after implementing a clean air plan in the Agglomeration Lausanne-Morges. Int. J. Hyg. Environ. Health 220, 829–839. https://doi.org/10.1016/j.ijheh.2017.03.012

Chan, C.K., Yao, X., 2008. Air pollution in mega cities in China. Atmos. Environ. 42, 1–42. https://doi.org/10.1016/j.atmosenv.2007.09.003

Charles, M., Ziv, G., Bohrer, G., Bakshi, B.R., 2020. Connecting air quality regulating ecosystem services with beneficiaries through quantitative serviceshed analysis. Ecosyst. Serv. 41, 101057. https://doi.org/10.1016/j.ecoser.2019.101057

Chen, K., Wang, M., Huang, C., Kinney, P.L., Anastas, P.T., 2020. Air pollution reduction and mortality benefit during the COVID-19 outbreak in China. Lancet Planet. Heal. 2019, 2019–2021. https://doi.org/10.1016/S2542-5196(20)30107-8

Chinazzi, M., Davis, J.T., Ajelli, M., Gioannini, C., Litvinova, M., Merler, S., Pastore y Piontti, A., Mu, K., Rossi, L., Sun, K., Viboud, C., Xiong, X., Yu, H., Halloran, M.E., Longini, I.M., Vespignani, A., 2020. The effect of travel restrictions on the spread of the 2019 novel coronavirus (COVID-19) outbreak. Science (80-.). 368, 395 LP – 400. https://doi.org/10.1126/science.aba9757

COMEAP, 2010. “The Mortality Effects of Long Term Exposure to Particulate AirPollution in the UK”, Report Produced by the Health Protection Agency for theCommittee on the Medical Effects of Air Pollutants, ISBN 978–0–85951–685–3,98 pp

Comberti, C., Thornton, T.F., Wyllie de Echeverria, V., Patterson, T., 2015. Ecosystem services or services to ecosystems? Valuing cultivation and reciprocal relationships between humans and ecosystems. Glob. Environ. Chang. 34, 247–262. https://doi.org/10.1016/j.gloenvcha.2015.07.007

Costanza, R., d’Arge, R., De Groot, R., Farber, S., Grasso, M., Hannon, B., Limburg, K., Naeem, S., O’neill, R. V, Paruelo, J., 1997. The value of the world’s ecosystem services and natural capital. Nature 387, 253–260.

Costanza, R., De Groot, R., Sutton, P., Van der Ploeg, S., Anderson, S.J., Kubiszewski, I., Farber, S., Turner, R.K., 2014. Changes in the global value of ecosystem services. Glob. Environ. Chang. 26, 152–158.

De Brouwer, E., Raimondi, D., Moreau, Y., 2020. Modeling the COVID-19 outbreaks and the effectiveness of the containment measures adopted across countries. medRxiv 2020.04.02.20046375. https://doi.org/10.1101/2020.04.02.20046375

De Carvalho, R.M., Szlafsztein, C.F., 2019. Urban vegetation loss and ecosystem services: The influence on climate regulation and noise and air pollution. Environ. Pollut. 245, 844–852. https://doi.org/10.1016/j.envpol.2018.10.114

Dutheil, F., Baker, J.S., Navel, V., 2020a. COVID-19 as a factor influencing air pollution? Environ. Pollut. 263, 2019–2021. https://doi.org/10.1016/j.envpol.2020.114466

Dutheil, F., Baker, J.S., Navel, V., 2020b. COVID-19 as a factor influencing air pollution? Environ. Pollut. 263, 114466. https://doi.org/10.1016/j.envpol.2020.114466

Etchie, T.O., Etchie, A.T., Adewuyi, G.O., Pillarisetti, A., Sivanesan, S., Krishnamurthi, K., Arora, N.K., 2018. The gains in life expectancy by ambient PM2.5 pollution reductions in localities in Nigeria. Environ. Pollut. 236, 146–157. https://doi.org/10.1016/j.envpol.2018.01.034

Escobedo, F.J., Wagner, J.E., Nowak, D.J., De la Maza, C.L., Rodriguez, M., Crane, D.E., 2008. Analysing the cost effectiveness of Santiago, Chile’s policy of using urban forests to improve air quality. J. Environ. Manage. 86, 148–157. https://doi.org/10.1016/j.jenvman.2006.11.029

Feng, L., Liao, W., 2016. Legislation, plans, and policies for prevention and control of air pollution in China: achievements, challenges, and improvements. J. Clean. Prod. 112, 1549–1549. https://doi.org/10.1016/j.jclepro.2015.08.013

Fisher, B., Turner, R.K., Morling, P., 2009. Defining and classifying ecosystem services for decision making. Ecol. Econ. 68, 643–653. https://doi.org/10.1016/j.ecolecon.2008.09.014

Gautam, S., 2020. The Influence of COVID-19 on Air Quality in India: A Boon or Inutile. Bull. Environ. Contam. Toxicol. 104, 724–726. https://doi.org/10.1007/s00128-020-02877-y

Gómez-Baggethun, E., Barton, D.N., 2013. Classifying and valuing ecosystem services for urban planning. Ecol. Econ. 86, 235–245. https://doi.org/10.1016/j.ecolecon.2012.08.019

Goscé, L., Johansson, A., 2018. Analysing the link between public transport use and airborne transmission: mobility and contagion in the London underground. Environ. Heal. 17, 84. https://doi.org/10.1186/s12940-018-0427-5

Griffin, D., Zhao, X., McLinden, C.A., Boersma, F., Bourassa, A., Dammers, E., Degenstein, D., Eskes, H., Fehr, L., Fioletov, V., Hayden, K., Kharol, S.K., Li, S.-M., Makar, P., Martin, R. V, Mihele, C., Mittermeier, R.L., Krotkov, N., Sneep, M., Lamsal, L.N., Linden, M. ter, Geffen, J. van, Veefkind, P., Wolde, M., 2019. High-Resolution Mapping of Nitrogen Dioxide With TROPOMI: First Results and Validation Over the Canadian Oil Sands. Geophys. Res. Lett. 46, 1049–1060. https://doi.org/10.1029/2018GL081095

Guanter, L., Aben, I., Tol, P., Krijger, J.M., Hollstein, A., Köhler, P., Damm, A., Joiner, J., Frankenberg, C., Landgraf, J., 2015. Potential of the TROPOspheric Monitoring Instrument (TROPOMI) onboard the Sentinel-5 Precursor for the monitoring of terrestrial chlorophyll fluorescence. Atmos. Meas. Tech. 8, 1337–1352. https://doi.org/10.5194/amt-8-1337-2015

Guerriero, C., Chatzidiakou, L., Cairns, J., Mumovic, D., 2016. The economic benefits of reducing the levels of nitrogen dioxide (NO2) near primary schools: The case of London. J. Environ. Manage. 181, 615–622. https://doi.org/10.1016/j.jenvman.2016.06.039

Guttikunda, S.K., Goel, R., Pant, P., 2014. Nature of air pollution, emission sources, and management in the Indian cities. Atmos. Environ. 95, 501–510. https://doi.org/10.1016/j.atmosenv.2014.07.006

He, G., Pan, Y., Tanaka, T., 2020. The short-term impacts of COVID-19 lockdown on urban air pollution in China. Nat. Sustain. https://doi.org/10.1038/s41893-020-0581-y

He, L., Zhang, S., Hu, J., Li, Z., Zheng, X., Cao, Y., Xu, G., Yan, M., Wu, Y., 2020. On-road emission measurements of reactive nitrogen compounds from heavy-duty diesel trucks in China. Environ. Pollut. 262, 114280. https://doi.org/10.1016/j.envpol.2020.114280

Hu, J., Ying, Q., Wang, Y., Zhang, H., 2015. Characterising multi-pollutant air pollution in China: Comparison of three air quality indices. Environ. Int. 84, 17–25. https://doi.org/10.1016/j.envint.2015.06.014

Jeanjean, A.P.R., Gallagher, J., Monks, P.S., Leigh, R.J., 2017. Ranking current and prospective NO2 pollution mitigation strategies: An environmental and economic modelling investigation in Oxford Street, London. Environ. Pollut. 225, 587–597. https://doi.org/10.1016/j.envpol.2017.03.027

Kanniah, K.D., Kamarul Zaman, N.A.F., Kaskaoutis, D.G., Latif, M.T., 2020. COVID-19’s impact on the atmospheric environment in the Southeast Asia region. Sci. Total Environ. 736, 139658. https://doi.org/10.1016/j.scitotenv.2020.139658

Kaplan, G., YİGİT AVDAN, Z., 2020. Space-Borne Air Pollution Observation From Sentinel-5P Tropomi: Relationship Between Pollutants, Geographical and Demographic Data. Int. J. Eng. Geosci. 130–137. https://doi.org/10.26833/ijeg.644089

Kerimray, A., Baimatova, N., Ibragimova, O.P., Bukenov, B., Kenessov, B., 2020. Since January 2020 Elsevier has created a COVID-19 resource centre with free information in English and Mandarin on the novel coronavirus COVID-19. The COVID-19 resource centre is hosted on Elsevier Connect, the company’s public news and information.

Kim Oanh, N.T., Upadhyay, N., Zhuang, Y.-H., Hao, Z.-P., Murthy, D.V.S., Lestari, P., Villarin, J.T., Chengchua, K., Co, H.X., Dung, N.T., Lindgren, E.S., 2006. Particulate air pollution in six Asian cities: Spatial and temporal distributions, and associated sources. Atmos. Environ. 40, 3367–3380.

Environ. https://doi.org/10.1016/j.atmosenv.2006.01.050

Kumar, P., Khare, M., Harrison, R.M., Bloss, W.J., Lewis, A., Coe, H., Morawska, L., 2015. New directions: Air pollution challenges for developing megacities like Delhi. Atmospheric Environment 122, 657–661.

Kumar, P., Andrade, M.F., Ynoue, R.Y., Fornaro, A., de Freitas, E.D., Martins Martins, J.L.D., Albuquerque, T., Zhang, Y., Morawska, L., 2016. New Directions: From biofuels to wood stoves: the modern and ancient air quality challenges in the megacity of São Paulo. Atmospheric Environment 140, 364–369.

Kumar, P., Druckman, A., Gallagher, J., Gatersleben, B., Allison, S., Eisenman, T.S., Hoang, U., Hama, S., Tiwari, A., Sharma, A., Abhijith, KV, Adlakha, D., McNabola, A., Astell-Burt, T., Feng, X., Skeldon, A.C., de Lusignan, S., Morawska, L., 2019. The Nexus between Air Pollution, Green Infrastructure and Human Health. Environment International 133,105181.

Kumar, P., Hama, S., Omidvarborna, H., Sharma, A., Sahani, J., Abhijith, K.V., Debele, S., Zavala-Reyes, J., Barwise, Y., Tiwari, A., 2020a. Temporary reduction in fine particulate matter due to ‘anthropogenic emissions switch-off’ during COVID-19 lockdown in Indian cities. Sustain. Cities Soc. 62, 102382. https://doi.org/10.1016/j.scs.2020.102382

Kumar, P, and Lidia M. “Could fighting airborne transmission be the next line of defence against COVID-19 spread?.” City and Environment Interactions (2020): 100033.

L., C.D., A., P.P., Perry, H., R., B.J., Aaron van D., V., M.R., J., V.P., Michael, J., S., G.M., Arden, P.C., Michael, B., D., B.R., Alain, R., Richard, M., T., B.R., 2015. Ambient PM2.5, O3, and NO2 Exposures and Associations with Mortality over 16 Years of Follow-Up in the Canadian Census Health and Environment Cohort (CanCHEC). Health Perspect. 123, 1180–1186. https://doi.org/10.1289/ehp.1409276

Lorente, A., Boersma, K.F., Eskes, H.J., Veefkind, J.P., van Geffen, J.H.G.M., de Zeeuw, M.B., Denier van der Gon, H.A.C., Beirle, S., Krol, M.C., 2019. Quantification of nitrogen oxides emissions from build-up of pollution over Paris with TROPOMI. Sci. Rep. 9, 20033. https://doi.org/10.1038/s41598-019-56428-5

MA (Millennium Ecosystem Assessment) (2005) Ecosystems and Human Well-being: Synthesis. Island Press, Washington DC

Mahato, S., Pal, S., Ghosh, K.G., 2020. Effect of lockdown amid COVID-19 pandemic on air quality of the megacity Delhi, India. Sci. Total Environ. 730, 139086. https://doi.org/10.1016/j.scitotenv.2020.139086

Matthews, H.S., Lave, L.B., 2000. Applications of Environmental Valuation for Determining Externality Costs. Environ. Sci. Technol. 34, 1390–1395. https://doi.org/10.1021/es9907313

Mayer, H., 1999. Air pollution in cities. Atmos. Environ. 33, 4029–4037. https://doi.org/10.1016/S1352-2310(99)00144-2

Monica, C., Ennio, C., Massimo, S., Claudia, G., Giovanna, B., Annunziata, F., Luigi, B., Angela, V.M., Patrizia, D.M., Achille, C., Sandra, M., Barbara, P., Sante, M., Lorenzo, S., Francesco, F., null, null, 2011. Short-Term Effects of Nitrogen Dioxide on Mortality and Susceptibility Factors in 10 Italian Cities: The EpiAir Study. Environ. Health Perspect. 119, 1233–1238. https://doi.org/10.1289/ehp.1002904

Muhammad, S., Long, X., Salman, M., 2020. COVID-19 pandemic and environmental pollution: A blessing in disguise? Sci. Total Environ. 728, 138820. https://doi.org/10.1016/j.scitotenv.2020.138820

Nowak, David J. 1994. Understanding the structure. Journal of Forestry. 92(10): 42–46.

Ogen, Y., 2020. Assessing nitrogen dioxide (NO2) levels as a contributing factor to coronavirus (COVID-19) fatality. Sci. Total Environ. 726, 138605. https://doi.org/10.1016/j.scitotenv.2020.138605

Ortiz, C., Linares, C., Carmona, R., Díaz, J., 2017. Evaluation of short-term mortality attributable to particulate matter pollution in Spain. Environ. Pollut. 224, 224–541. https://doi.org/10.1016/j.envpol.2017.02.037

Otmani, A., Benchrif, A., Tahri, M., Bounakhla, M., Chakir, E.M., El Bouch, M., Krombi, M., 2020. Impact of Covid-19 lockdown on PM10, SO2 and NO2 concentrations in Salé City (Morocco). Sci. Total Environ. 735, 139541. https://doi.org/10.1016/j.scitotenv.2020.139541

Pandeya, B., Buytaert, W., Zulkafli, Z., Karpouzoglou, T., Mao, F., Hannah, D.M., 2016. A comparative analysis of ecosystem services valuation approaches for application at the local scale and in data scarce regions. Ecosyst. Serv. 22, 250–259. https://doi.org/10.1016/j.ecoser.2016.10.015

Pilla, F., Broderick, B., 2015. A GIS model for personal exposure to PM10 for Dublin commuters. Sustain. Cities Soc. 15, 1–10. https://doi.org/10.1016/j.scs.2014.10.005

Potschin, M., Haines-Young, R., 2013. Landscapes, sustainability and the place-based analysis of ecosystem services. Landsc. Ecol. 28, 1053–1065. https://doi.org/10.1007/s10980-012-9756-x

Rai, A.C., Kumar, P., Pilla, F., Skouloudis, A.N., Di Sabatino, S., Ratti, C., Yasar, A., Rickerby, D., 2017. End-user perspective of low-cost sensors for outdoor air pollution monitoring. Sci. Total Environ. 607–608, 691–705. https://doi.org/10.1016/j.scitotenv.2017.06.266

Rohde, R.A., Muller, R.A., 2015. Air Pollution in China: Mapping of Concentrations and Sources. PLoS One 10, e0135749.

Sahu, S.K., Kota, S.H., 2017. Significance of PM2.5 air quality at the Indian capital. Aerosol Air Qual. Res. 17, 588–597. https://doi.org/10.4209/aaqr.2016.06.0262

Sannigrahi, S., Bhatt, S., Rahmat, S., Paul, S.K., Sen, S., 2018. Estimating global ecosystem service values and its response to land surface dynamics during 1995–2015. J. Environ. Manage. 223. https://doi.org/10.1016/j.jenvman.2018.05.091

Sannigrahi, S., Chakraborti, S., Joshi, P.K., Keesstra, S., Sen, S., Paul, S.K., Kreuter, U., Sutton, P.C., Jha, S., Dang, K.B., 2019a. Ecosystem service value assessment of a natural reserve region for strengthening protection and conservation. J. Environ. Manage. 244, 208–227. https://doi.org/10.1016/j.jenvman.2019.04.095

Sannigrahi, S., Pilla, F., Basu, B., Basu, A.S. and Molter, A., 2020a. Examining the association between socio-demographic composition and COVID-19 fatalities in the European region using spatial regression approach. Sustainable Cities and Society, p.102418.

Sannigrahi, S., Zhang, Q., Joshi, P.K., Sutton, P.C., Keesstra, S., Roy, P.S., Pilla, F., Basu, B., Wang, Y., Jha, S., 2020b. Examining effects of climate change and land use dynamic on biophysical and economic values of ecosystem services of a natural reserve region. J. Clean. Prod. 257, 120424.

Sannigrahi, S., Zhang, Q., Pilla, F., Joshi, P.K., Basu, B., Keesstra, S., Roy, P.S., Wang, Y., Sutton, P.C., Chakraborti, S., 2020c. Responses of ecosystem services to natural and anthropogenic forcings: A spatial regression based assessment in the world’s largest mangrove ecosystem. Sci. Total Environ. 715, 137004.

Sasidharan, M., Singh, A., Torbaghan, M.E., Parlikad, A.K., 2020. A vulnerability-based approach to human-mobility reduction for countering COVID-19 transmission in London while considering local air quality. Sci. Total Environ. 741, 140515. https://doi.org/10.1016/j.scitotenv.2020.140515

Schirpke, U., Scolozzi, R., De Marco, C., Tappeiner, U., 2014. Mapping beneficiaries of ecosystem services flows from Natura 2000 sites. Ecosyst. Serv. 9, 170–179. https://doi.org/10.1016/j.ecoser.2014.06.003

Sharma, S., Zhang, M., Anshika Gao, J., Zhang, H., Kota, S.H., 2020. Effect of restricted emissions during COVID-19 on air quality in India. Sci. Total Environ. 728, 138878. https://doi.org/10.1016/j.scitotenv.2020.138878

Shehzad, K., Sarfraz, M., Shah, S.G.M., 2020. The impact of COVID-19 as a necessary evil on air pollution in India during the lockdown. Environ. Pollut. 266, 115080. https://doi.org/10.1016/j.envpol.2020.115080

Shikwambana, L., Mhangara, P., Mbatha, N., 2020. Trend analysis and first time observations of sulphur dioxide and nitrogen dioxide in South Africa using TROPOMI/Sentinel-5 P data. Int. J. Appl. Earth Obs. Geoinf. 91, 102130. https://doi.org/10.1016/j.jag.2020.102130

Sicard, P., De Marco, A., Agathokleous, E., Feng, Z., Xu, X., Paoletti, E., Rodriguez, J.J.D., Calatayud, V., 2020. Amplified ozone pollution in cities during the COVID-19 lockdown. Sci. Total Environ. 735. https://doi.org/10.1016/j.scitotenv.2020.139542

Spangenberg, J.H., von Haaren, C., Settele, J., 2014. The ecosystem service cascade: Further developing the metaphor. Integrating societal processes to accommodate social processes and planning, and the case of bioenergy. Ecol. Econ. 104, 22–32. https://doi.org/10.1016/j.ecolecon.2014.04.025

Theys, N., Hedelt, P., De Smedt, I., Lerot, C., Yu, H., Vlietinck, J., Pedergnana, M., Arellano, S., Galle, B., Fernandez, D., Carlito, C.J.M., Barrington, C., Taisne, B., Delgado-Granados, H., Loyola, D., Van Roozendael, M., 2019. Global monitoring of volcanic SO2 degassing with unprecedented resolution from TROPOMI onboard Sentinel-5 Precursor. Sci. Rep. 9, 2643. https://doi.org/10.1038/s41598-019-39279-y

Troko, J., Myles, P., Gibson, J., Hashim, A., Enstone, J., Kingdon, S., Packham, C., Amin, S., Hayward, A., Van-Tam, J.N., 2011. Is public transport a risk factor for acute respiratory infection? BMC Infect. Dis. 11, 16. https://doi.org/10.1186/1471-2334-11-16

Valade, S., Ley, A., Massimetti, F., D’Hondt, O., Laiolo, M., Coppola, D., Loibl, D., Hellwich, O., Walter, T.R., 2019. Towards global volcano monitoring using multisensor sentinel missions and artificial intelligence: The MOUNTS monitoring system. Remote Sens. 11, 1–31. https://doi.org/10.3390/rs11131528

Veefkind, J.P., Aben, I., McMullan, K., Förster, H., de Vries, J., Otter, G., Claas, J., Eskes, H.J., de Haan, J.F., Kleipool, Q., van Weele, M., Hasekamp, O., Hoogeveen, R., Landgraf, J., Snel, R., Tol, P., Ingmann, P., Voors, R., Kruizinga, B., Vink, R., Visser, H., Levelt, P.F., 2012. TROPOMI on the ESA Sentinel-5 Precursor: A GMES mission for global observations of the atmospheric composition for climate, air quality and ozone layer applications. Remote Sens. Environ. 120, 70–83. https://doi.org/10.1016/j.rse.2011.09.027

Venter, Z.S., Aunan, K., Chowdhury, S., Lelieveld, J., 2020. COVID-19 lockdowns cause global air pollution declines with implications for public health risk. medRxiv 2020.04.10.20060673. https://doi.org/10.1101/2020.04.10.20060673

Viscusi, W.K., Masterman, C.J., 2017. Income Elasticities and Global Values of a Statistical Life. J. Benefit-Cost Anal. 8, 226–250. https://doi.org/10.1017/bca.2017.12

Wang, S., Hao, J., 2012. Air quality management in China: Issues, challenges, and options. J. Environ. Sci. 24, 2–13. https://doi.org/10.1016/S1001-0742(11)60724-9

Woodcock, C. E., Allen, R., Anderson, M., Belward, A., Bindschadler, R., Cohen, W., … & Nemani, R. (2008). Free access to Landsat imagery. SCIENCE VOL 320: 1011.

Zhang, H., Wang, Shuxiao, Hao, J., Wang, X., Wang, Shulan, Chai, F., Li, M., 2016. Air pollution and control action in Beijing. J. Clean. Prod. 112, 1519–1527. https://doi.org/10.1016/j.jclepro.2015.04.092

Zhang, Q., Song, C. and Chen, X., 2018. Effects of China’s payment for ecosystem services programs on cropland abandonment: A case study in Tiantangzhai Township, Anhui, China. Land use policy, 73, pp.239–248.

Zhang, Q., Wang, Y., Tao, S., Bilsborrow, R.E., Qiu, T., Liu, C., Sannigrahi, S., Li, Q. and Song, C., 2020. Divergent socioeconomic-ecological outcomes of China’s conversion of cropland to forest program in the subtropical mountainous area and the semi-arid Loess Plateau. Ecosystem Services, 45, p.101167.

Zheng, Z., Yang, Z., Wu, Z., Marinello, F., 2019. Spatial variation of NO2 and its impact factors in China: An application of sentinel-5P products. Remote Sens. 11, 1–24. https://doi.org/10.3390/rs11161939

Zhu, Y., Xie, J., Huang, F., Cao, L., 2020. Association between short-term exposure to air pollution and COVID-19 infection: Evidence from China. Sci. Total Environ. 727, 138704. https://doi.org/10.1016/j.scitotenv.2020.138704

